# *cfTrack*: Exome-wide mutation analysis of cell-free DNA to simultaneously monitor the full spectrum of cancer treatment outcomes: MRD, recurrence, and evolution

**DOI:** 10.1101/2021.11.07.21265999

**Authors:** Shuo Li, Weihua Zeng, Xiaohui Ni, Yonggang Zhou, Mary L. Stackpole, Zorawar S. Noor, Zuyang Yuan, Adam Neal, Sanaz Memarzadeh, Edward B. Garon, Steven M. Dubinett, Wenyuan Li, Xianghong Jasmine Zhou

## Abstract

**Purpose:** Cell-free DNA (cfDNA) offers a non-invasive approach to monitor cancer. Here we develop a method using whole-exome sequencing (WES) of cfDNA for simultaneously monitoring the full spectrum of cancer treatment outcomes, including MRD, recurrence, evolution, and second primary cancer.

**Experimental Design:** Three simulation datasets were generated from 26 cancer patients to benchmark the detection performance of MRD/recurrence and second primary cancers. For further validation, cfDNA samples (n=76) from cancer patients (n=35) with six different cancer types were used for validating the performance of cancer monitoring during various treatments.

**Results:** We present a cfDNA-based cancer monitoring method, named *cfTrack*. Taking advantage of the broad genome coverage of WES data, *cfTrack* can sensitively detect MRD and cancer recurrence by integrating signals across the known clonal tumor mutations of a patient. In addition, *cfTrack* detects tumor evolution and second primary cancers by *de novo* identifying emerging tumor mutations. A series of machine learning and statistical denoising techniques are applied to enhance the detection power. On the simulation data, *cfTrack* achieved an average AUC of 99% on the validation dataset and 100% on the independent dataset in detecting recurrence in samples with tumor fraction ≥0.05%. In addition, *cfTrack* yielded an average AUC of 88% in detecting second primary cancers in samples with tumor fraction ≥0.2%. On real data, *cfTrack* accurately monitors tumor evolution during treatment, which cannot be accomplished by previous methods.

**Conclusion:** Our results demonstrated that *cfTrack* can sensitively and specifically monitor the full spectrum of cancer treatment outcomes using exome-wide mutation analysis of cfDNA.

**Translational Relevance:** Continuous cancer monitoring is clinically necessary for cancer patients to detect minimal residual disease (MRD), recurrence, and progression, allowing for early intervention and therapy adjustment. Cell-free DNA (cfDNA) in blood has become an appealing option due to its non-invasiveness. Until now, cfDNA-based cancer monitoring methods have been focused on deep sequencing at a few known mutations, which are however insufficient when tumors evolve or new tumors emerge. We present the method, cfTrack, which for the first time uses whole-exome sequencing (WES) of cfDNA to track the full range of cancer treatment outcomes, including MRD, recurrence, evolution, and second primary cancer. We demonstrate that, even with very low tumor fractions, cfTrack achieves sensitive and specific monitoring of tumor MRD/recurrence/evolution based on both simulation data and a cohort of cancer patients. These findings demonstrate the clinical utility of cfTrack.

## Introduction

Despite the rapid development of cancer treatments, a large fraction of patients experiences recurrence, resistance, or progression of cancer during or after treatment [1]. Even after the surgical removal of tumors, a patient can still have minimal residual disease (MRD), which is associated with an increased likelihood of recurrence [2]. Thus, cancer patients need continuous monitoring in order to detect MRD, recurrence, and progression, thereby facilitating the early intervention and the therapy adjustment [2][3]. Although cancer monitoring is clinically important, the sequential sampling of tumor tissue from the patient poses a significant challenge. In this context, liquid biopsy is an attractive option, especially the usage of **c**ell-**f**ree **DNA** (**cfDNA**) in blood. Blood samples can be obtained noninvasively for continuous monitoring, and the tumor-derived DNA fragments in cfDNA can provide comprehensive genetic profiling even of heterogeneous tumors [4].

However, a major challenge associated with cfDNA-based cancer monitoring is the low tumor content. In cancer patients receiving treatment or with MRD, the fraction of tumor DNA in a cfDNA sample can be as low as 0.1% [5]. Previous studies on cancer monitoring in plasma have used deep sequencing on a small mutation panel to discover the weak tumor signal [2][5][6][7][8]. However, these methods have several crucial limitations: (1) the high cost of deep sequencing restricts the panels to a small number of known mutations (either common cancer mutations or mutations selected from the pre-treatment tumor sample of a specific patient); (2) personalized panels usually require a labor-intensive experimental design; (3) panel-based monitoring cannot detect emerging tumors with a different mutation profile, e.g., a second primary cancer, yet approximately 30% patients develop a second primary cancer [9], driven by *de novo* tumor mutations; and (4) panels usually set detection thresholds by studying a cohort of non-cancer individuals, which exposes the test to systemic bias from inter-individual variations and inter-experimental differences. Recently, two studies [10][11] presented cancer monitoring methods using whole-genome sequencing, but they do not yet address limitations 3 and 4 mentioned above, and focus on mutations seen in pre-treatment tumor samples. In addition, the high cost of whole-genome sequencing limits the clinical applications of those two methods.

In this study, we describe a new cancer monitoring approach, named *cfTrack*, based on the cfDNA whole-exome sequencing (WES). *cfTrack* addresses all the aforementioned limitations of existing methods. Specifically, not only can it monitor the pre-existing cancer (i.e., the original, primary cancer) to detect recurrence or MRD, but it can also monitor tumor evolution by detecting cancer progression or the emergence of a second primary cancer. To monitor the pre-existing cancer, *cfTrack* (1) uses exome-wide somatic mutations collected in pre-treatment samples (solid tumor or blood samples) to provide a robust statistical index, then (2) models sample-specific background noise in the cfDNA sequencing data to provide an unbiased detection threshold for each patient. To monitor tumor evolution, *cfTrack* performs detection of exome-wide *de novo* tumor mutations in the post-treatment plasma samples, using our recently developed *cfSNV* method [12]. With exome-wide sequencing and comprehensive analysis of mutations, *cfTrack* can sensitively identify these previously undiagnosed patients with second primary cancers, comprehensively describe their tumor status, and enable early intervention and personalization of treatment. Using both simulation data and a cohort of cancer patients (n = 35, 18 prostate cancer, 8 lung cancer, 4 ovarian cancer, 3 glioma, 1 bladder cancer, and 1 germ cell cancer), we show that *cfTrack* achieves sensitive and specific monitoring of tumor MRD/recurrence and evolution from cfDNA, even with very low tumor fractions. These results demonstrate that *cfTrack* enables full-spectrum monitoring of cancer treatment outcomes.

## Material and Methods

### Data collection

We collected WES data from four public datasets. We collected data of 18 metastatic cancer patients from Adalsteinsson et al. [17] under dbGaP accession code phs001417.v1.p1. Each patient’s data includes a WBC sample, a tumor biopsy sample, and two plasma samples. We also collected WES data of 3 cancer patients (1 bladder cancer, 1 prostate cancer, and 1 germ cell cancer) from Tsui et al. [27] under dbGaP accession code phs002290 and WES data of 3 glioma patients under SRA accession code SRP268702. Each patient has a WBC sample, a solid tumor sample, and a plasma sample. We collected WES data of 17 prostate cancer patients from Ramesh et al. [26] under SRA accession code SRP260849. All patients have one WBC sample; 8 of the 17 patients have a solid tumor sample (metastatic site); 5 (7, 2, 2, and 1) patients have 1 (2, 3, 4, and 5 respectively) plasma sample collected at different time points. We also collected samples from 8 NSCLC patients and 4 ovarian cancer patients and generated our own WES data as described below. For all 8 NSCLC patients, a tumor biopsy sample, a WBC sample, and three plasma samples were collected. For all 4 ovarian cancer patients, a WBC sample and two serum samples were collected. In addition, for the ovarian cancer patient OV4, who underwent surgical resection at the first blood collection, we collected the patient’s tumor tissue sample. For all sources, only one WBC sample (or its WES data) was collected for each cancer patient.

### Human subjects

We collected blood samples, tumor samples, and WBC samples from 8 NSCLC patients from KEYNOTE-001 [30] and KEYNOTE-010 [31], who all provided informed consent for research use. The blood and tissue collection protocols were described in the full protocol of KEYNOTE-001 and KEYNOTE-010. The project was approved by the Institutional Review Board (IRB) of University of California, Los Angeles (IRB# 12-001891, IRB# 11-003066, and IRB# 13-00394) and was conducted in accordance with the Belmont Report. We also collected samples from 4 ovarian cancer patients. Serum was harvested from whole blood by centrifugation (400xg, 15’) and immediately flash frozen. PBMCs were harvested from whole blood collected in a blue top phlebotomy tube with sodium citrate, centrifuged (400xg, 15’), and aliquoted from the buffy coat before being immediately flash frozen. Portions of solid tumor from the operating room were brought back to the lab and flash frozen. Clinical information from consenting patients was obtained from medical records. Longitudinally collected clinical specimens from ovarian cancer patients were obtained using IRB-approved protocols (IRB# 10-000727) and were studied in accordance with the Belmont Report. All patients provided written informed consent.

### Genomic DNA WES library construction

For the 8 NSCLC patients, the WBC samples underwent multiplexed paired-end WES to a target depth of 100-150x on HiSeq 2000/3000 (Illumina, San Diego, CA) performed by the UCLA Technology Center for Genomics & Bioinformatics. Macrodissection was not performed. DNA isolation was performed with DNeasy Blood & Tissue Kit (Qiagen, Germany); exon capture and library preparation for the 8 NSCLC patients (WBC samples) used the KAPA HyperPrep Kit and Nimblegen SeqCap EZ Human Exome Library v3.0 (Roche, Switzerland) before the final step of 2×150bp paired-end sequencing by Genewiz (South Plainfield, NJ). For the 4 ovarian cancer patients, the WBC gDNA and the tumor tissue gDNA isolation were performed with DNeasy Blood & Tissue Kit (Qiagen) and sonicated by Covaris system (Woburn, MA). Ampure XP beads (Beckman-Coulter, Atlanta, GA) size selection was further performed to enrich the fragments between 100 and 250bp. In brief, 0.9 volume of beads were first added to the fragmented gDNA samples. After incubation, the supernatant was transferred to a new tube and an additional 1.1 volume of beads were added. After 80% ethanol wash, the size-selected gDNA was eluted from the beads. The gDNA WES library was constructed with the SureSelect XT HS kit from Agilent Technologies (Santa Clara, CA) according to the manufacturer’s protocol. No molecular barcodes were used in the sequencing libraries. In brief, 100ng of gDNA was used as input material. After end repair/dA-tailing of cfDNA, the adaptor was ligated. The ligation product was purified with Ampure XP beads and the adaptor-ligated library was amplified with index primer in 8-cycle PCR. The amplified library was purified again with Ampure XP beads, and the amount of amplified DNA was measured using the Qubit 1xdsDNA HS assay kit (ThermoFisher, Waltham, MA). 1000 ng of DNA sample was hybridized to the capture library and pulled down by streptavidin-coated beads (ThermoFisher). After washing the beads, the DNA library captured on the beads was re-amplified with 9-cycles of PCR. The final libraries were purified by Ampure XP beads. The library concentration was measured by Qubit. The library quality check was further performed with Agilent Bioanalyzer before the final step of 2×150bp paired-end sequencing by Genewiz (South Plainfield, NJ).

### Plasma cfDNA WES library construction

For each of the 8 NSCLC patients, venipuncture was performed by trained phlebotomists such as nurses or medical assistants. Blood tubes were centrifuged at 1,800g for 20 min at room temperature and plasma supernatant was isolated within 2 hours of collection. Samples were stored at -80ºC until use. Then, cfDNA was extracted from their plasma samples using the QIAamp circulating nucleic acid kit from QIAGEN (Germantown, MD). The cfDNA WES library was constructed with the SureSelect XT HS kit from Agilent Technologies (Santa Clara, CA) according to the manufacturer’s protocol. No molecular barcodes were used in the sequencing libraries. In brief, 10ng of cfDNA was used as input material. After end repair/dA-tailing of cfDNA, the adaptor was ligated. The ligation product was purified with Ampure XP beads (Beckman-Coulter, Atlanta, GA) and the adaptor-ligated library was amplified with index primer in 10-cycle PCR. The amplified library was purified again with Ampure XP beads, and the amount of amplified DNA was measured using the Qubit 1xdsDNA HS assay kit (ThermoFisher, Waltham, MA). 700-1000 ng of DNA sample was hybridized to the capture library and pulled down by streptavidin-coated beads. After washing the beads, the DNA library captured on the beads was re-amplified with 10-cycles of PCR. The final libraries were purified by Ampure XP beads. The library concentration was measured by Qubit, and the quality was further examined with Agilent Bioanalyzer before the final step of 2×150bp paired-end sequencing by Genewiz (South Plainfield, NJ), at an average depth of 200x.

### Serum cfDNA WES library construction

For the serum samples from the four ovarian cancer patients, cfDNA was extracted by QIAamp circulating nucleic acid kit (QIAGEN). Ampure XP beads size selection was further performed to eliminate gDNA contamination. In brief, 0.5 volume of beads were first added to the cfDNA samples. After incubation, the supernatant was transferred to a new tube and an additional 2.0 volume of beads were added. After 80% ethanol wash, cfDNA was eluted from the beads. FA assays (Agilent Technologies) were performed to rule out the contamination of gDNA in the size selected samples. The cfDNA WES library was constructed with the SureSelect XT HS kit from Agilent Technologies (Santa Clara, CA) according to the manufacturer’s protocol. No molecular barcodes were used in the sequencing libraries. In brief, 5-20ng of gDNA was used as input material. After end repair/dA-tailing of cfDNA, the adaptor was ligated. The ligation product was purified with Ampure XP beads and the adaptor-ligated library was amplified with index primer in 11 cycles (for 10-20ng cfDNA input) or 12 cycles (for cfDNA less than 10ng). The amplified library was purified again with Ampure XP beads, and the amount of amplified DNA was measured using the Qubit 1xdsDNA HS assay kit (ThermoFisher, Waltham, MA). 1000 ng of DNA sample was hybridized to the capture library and pulled down by streptavidin-coated beads (ThermoFisher). After washing the beads, the DNA library captured on the beads was re-amplified with 9-cycles of PCR. The final libraries were purified by Ampure XP beads. The library concentration was measured by Qubit. The library quality check was further performed with Agilent Bioanalyzer before the final step of 2×150bp paired-end sequencing by Genewiz (South Plainfield, NJ).

### Data preprocessing

Both genomic DNA sequencing data and cfDNA sequencing data were preprocessed using the same procedure. Raw sequencing data (FASTQ files) were aligned to the hg19 reference genome by *bwa mem* [32] and sorted by *samtools* [33]. Then, duplicated reads from PCR amplification were identified and removed by *picard tools MarkDuplicates* [34]. After this step, read group information was added to the bam file using *picard tools AddOrReplaceReadGroups*, and reads were realigned around indels using *GATK RealignerTargetCreator* and *IndelRealigner* [35][36]. After realignment, base quality scores were recalibrated using *GATK BaseRecalibrator* and *PrintReads*. All tools in the data preprocessing pipeline were used with their default settings. After data preprocessing, the resulting bam files were used as inputs for mutation detection and MRD detection.

### Predicting MRD/recurrence in the post-treatment samples using somatic mutations detected from the pre-treatment samples

We predict the presence of MRD/recurrence by tracking the cfDNA fragments containing tumor-derived somatic mutations (i.e., tumor-derived cfDNA fragments). Due to the low tumor fraction in the plasma samples from patients with MRD, we integrate all clonal mutations in the exome to enhance the tumor signal (see **Identification of clonal mutations in pre-treatment samples**, Figure 1b (1)). To limit the accumulation of sequencing errors during integration, we employ a machine learning model (see **Machine learning model for suppressing sequencing errors**, Figure 1b (2)) that can accurately distinguish reads with sequencing errors from true mutations. Then, the level of tumor-derived cfDNA fragments is compared with a background noise distribution generated from the same plasma sample by a permutation test (see **Identification of mutations and CHIP positions** and **Building background noise distribution using random genomic locations**, Figure 1b (3)). If the tumor-derived cfDNA fragments are significantly more abundant than the background noise in the sample (*p*-value <= 0.05), the patient is predicted as having MRD/recurrence. If no MRD/recurrence is detected, the post-treatment sample is subsequently examined for the presence of second primary cancers (see **Detection of a second primary cancer**, Figure 1b (4)).

**Figure 1.**
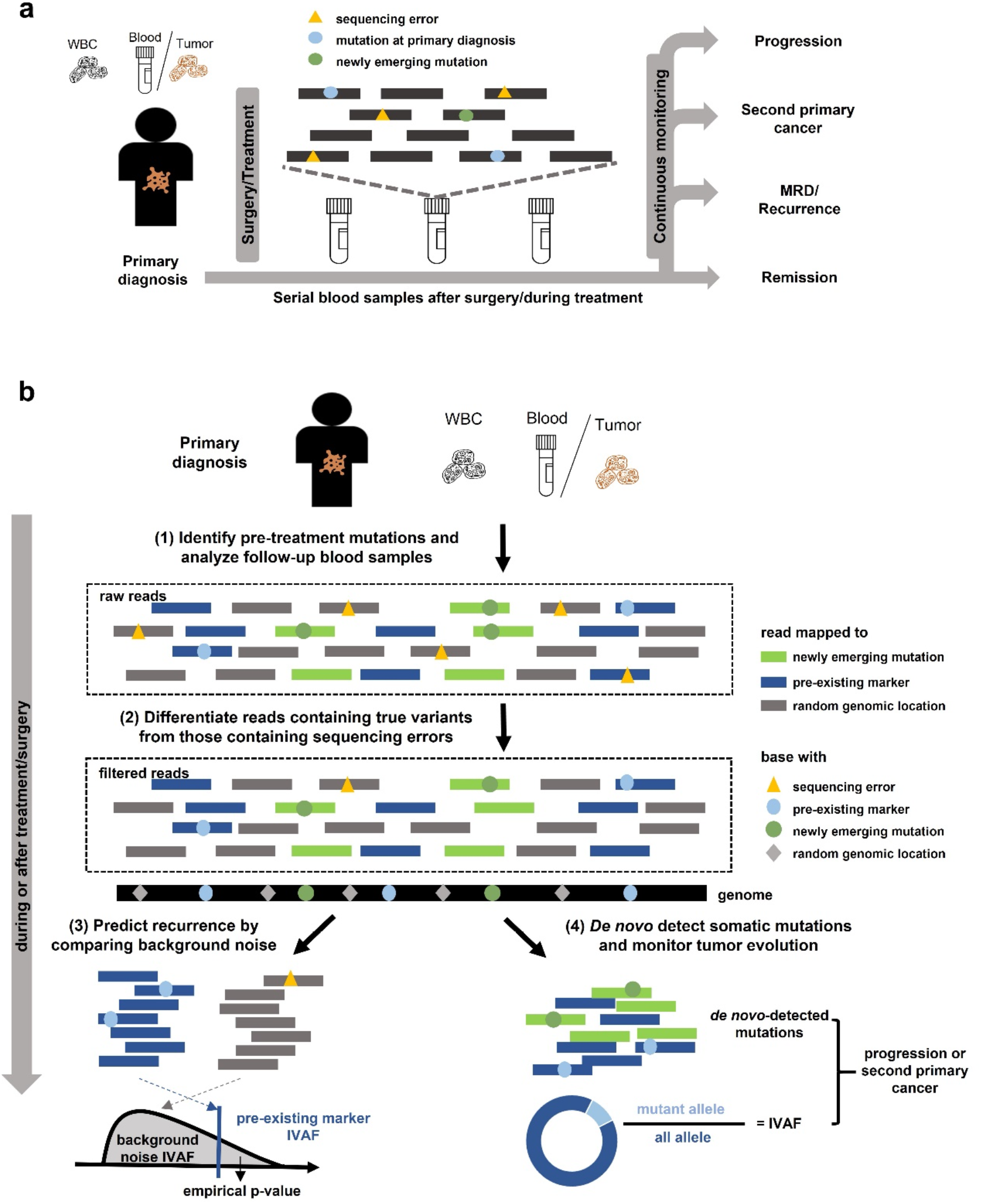
Cancer monitoring in plasma samples by tracking pre-existing tumor mutations and newly emerging tumor mutations. **(a)** Illustration of the sample collection for cfDNA-based cancer monitoring. Prior to surgery or therapy, a plasma or tumor sample and a white blood cell (WBC) sample are collected to generate the pre-existing tumor profile. Serial blood samples are collected to detect MRD/recurrence and monitor tumor evolution after treatment. **(b)** Illustration of the method workflow. In the pre-treatment samples, clonal tumor mutations are identified for tumor tracking in the post-treatment samples. Given a post-treatment plasma sample, the tumor fraction is calculated from the pre-existing clonal tumor mutations and compared to a sample-specific background distribution. The empirical p-value of the tumor fraction is used to predict MRD/recurrence. Furthermore, *de novo* somatic mutations are detected using *cfSNV* between the post-treatment plasma and WBC samples. A second primary cancer is predicted by a logistic regression model that accounts for both the amount of *de novo* mutations and the corresponding tumor fraction.

### Calculation of the Integrated Variant Allele Frequency (IVAF)

To quantify tumor DNA across multiple loci, we calculate the Integrated Variant Allele Frequency (IVAF) as the number of sequencing read pairs classified by the model as containing true mutations divided by the total number of read pairs. Both the numerator and the denominator are summed over all loci identified as clonal mutations in the pre-treatment sample. The IVAF indicates the fraction of high-confidence tumor DNA in all cfDNA fragments, so it is treated as the estimated tumor fraction in this study.

### Identification of clonal mutations in pre-treatment samples

The presence of tumor-derived somatic mutations in cfDNA is usually treated as a reliable marker to confirm the presence of cancer. However, not all tumor-derived somatic mutations are equally effective, because subclonal mutations have a lower observed allele frequency than clonal mutations [5]. To overcome the challenge of low tumor content in the plasma samples of patients with MRD, we integrate tumor-derived somatic mutations over a wide range of the genome (e.g., the whole exome data obtained by WES). The integration accumulates not only tumor-derived signals but also sequencing errors. Therefore, we used clonal somatic mutations, called from the pre-treatment plasma sample or the pre-treatment tumor sample, as tumor markers. To make this selection, first tumor-derived somatic mutations are detected using *cfSNV* [12] from the pre-treatment plasma sample; if only the pre-treatment tumor sample is available, tumor-derived mutations are the common mutations detected by Strelka2 [37] somatic and MuTect [38] from the pre-treatment tumor sample. The detected mutations are removed if there is at least one variant supporting read in the matched WBC sample. A mutation is considered clonal, and hence retained in the final marker list, if its VAF is > 25% of the average of the five highest VAFs in the sample [39]. We require a minimum of 30 markers from the pre-treatment plasma sample to obtain a robust prediction. If there are fewer than 30 clonal mutations, subclonal mutations with the highest VAFs will be included.

### Identification of mutations and Clonal Hematopoiesis of Indeterminate Potential (CHIP) positions

To accurately estimate the background noise in a sequencing experiment, it is essential to remove the interference from non-reference alleles at the germline mutations, somatic mutations, and CHIP positions. To estimate the background, we identify germline mutations in the pre-treatment plasma sample and the matched WBC sample from the same patient using *GATK HaplotypeCaller* and *Strelka2 Germline* with the default settings. *GATK HaplotypeCaller* is applied to the plasma sample and the WBC sample individually; *Strelka2 Germline* is applied to the plasma-WBC sample pair. Somatic mutations are detected in the plasma sample and the matched WBC sample using *cfSNV* under default settings. The CHIP positions are identified from pileup files generated using *samtools mpileup*. If a non-mutated position has >= 3 variant supporting reads or a VAF > 1% in the matched WBC sample, it is regarded as a CHIP position. The selection of these parameters has little impact on the performance (Supplementary Figure 4a-b). All the identified germline mutations, somatic mutations and CHIP positions are excluded in the step of building the background noise distribution.

### Building a background noise distribution using random genomic locations

The simple presence of variant supporting reads at tumor-derived somatic mutations is not enough to determine the presence of MRD/recurrence, because they could be caused by sequencing errors. Therefore, to quantify the sequencing error frequency, we build a background noise distribution from the same plasma sample that we use to monitor MRD/recurrence. Unlike using a panel of normal samples from other sources, this approach avoids potential biases from inter-individual and inter-experimental differences. A background noise distribution is generated for the clonal tumor mutations that were identified from the pre-treatment sample for MRD/recurrence monitoring. For a set of *n* clonal tumor mutations, *n* positions are randomly selected from the targeted genomic region (e.g., the whole exome), excluding known mutations and CHIP positions. Ideally, all read pairs with non-reference alleles at these *n* positions are from sequencing errors, so the observed frequency of these reads represents the background noise level. The sequencing read pairs containing non-reference alleles at these *n* positions are extracted and input into the sequencing noise suppression model. The observed frequency of a non-reference allele (i.e., its integrated variant allele frequency) is calculated as the number of sequencing read pairs classified by the model as containing true mutations, divided by the total number of read pairs aligned to the *n* positions. We repeat the random sampling of *n* positions and calculate the observed frequency of non-reference alleles *K* times. Finally, the background noise distribution is built from the *K* observed frequencies of non-reference alleles at random *n* positions. By comparing the tumor fraction *θ* at the clonal mutations with the background noise distribution, an empirical *p*-value can be calculated as the rank of *θ* among the *K* background IVAFs (in a decreasing order). If the empirical *p*-value is <= 0.05, the patient is regarded as having MRD/recurrence. Based on our simulation, only minor differences in the detection threshold occur when K is set to 100, 500, or 1000. Therefore, in our simulation, we set K to 100.

### Machine learning model for suppressing sequencing errors

Although weak tumor signals in plasma samples can be amplified by integrating the variant supporting reads across a large genomic region, sequencing errors can also accumulate and possibly confound the tumor signal. Moreover, because of the low fraction of tumor DNA, the variant supporting reads at a single mutation are not sufficient to provide a robust and accurate estimation of site-level statistics (e.g., strand bias and average base quality) for error removal. Therefore, we developed a machine learning filter to eliminate reads with sequencing errors (Supplementary Figure 5). Specifically, for a group of genomic positions (tumor mutations or random positions), we classify the variant supporting reads with a random forest model to distinguish sequencing errors from true variants. Since all data in this study were generated from paired-end sequencing, in the following section, we detail the model for paired-end reads, but the principle can also be applied to single-end reads. With paired-end sequencing data, there are two types of read pairs with regards to a specific mutation site: one (non-overlapping read pair) covers the mutation site by one of its read mates, the other (overlapping read pair) covers the mutation site by both of its read mates (Supplementary Figure 5a). The overlapping read pair can provide two readouts of the mutation site on the DNA fragment in the sequencing library, but the non-overlapping read pair can only provide one readout. This means that the overlapping read pair naturally contains more information about the mutation site than the non-overlapping read pair, and the two readouts can serve as validation for each other. Therefore, we trained two independent random forest models to fully utilize the information in the non-overlapping read pair and the overlapping read pair. Please note that the random forest models in *cfTrack* classify sequencing errors and true variants in every read pair, i.e. read-level error suppression. It is different from the empirical variant score model in Strelka and the variant quality score model in GATK, which rely on site-level statistics (such as averaged base quality in all reads) to classify sequencing errors and true variants.

To train the random forest model, we used WES data from 18 patients: 12 with metastatic breast cancer (MBC) and 6 with metastatic prostate cancer (CRPC) [17](Supplementary Figure 5b). Each patient had four samples sequenced: two plasma samples (collected at two different time points), a WBC sample, and a tumor biopsy sample. We use the supporting cfDNA read pairs at known mutation (error) sites as the training data. The known mutation sites include both germline and somatic mutation sites, where germline mutations are required to be detected in all four samples using *Strelka2 germline*, and somatic mutations are required to be detected from both the cfDNA-WBC pairs (cfDNA data vs. WBC data) and the tumor-WBC pair (tumor data vs. WBC data) using *Strelka2 somatic* and *MuTect*. Error sites are defined as sufficiently covered sites (> 150x) with at most two high-quality non-reference read (base quality ≥ 20 and mapping quality ≥ 40) in only one of the four datasets. All high-quality labeled read pairs (base quality ≥ 30 and mapping quality ≥ 40) were extracted from raw cfDNA data using *picard tools FilterSamReads*. Multiple read pairs may be extracted covering the same mutation site, but these read pairs are similar and might cause redundancy in the training and testing data. Therefore, we solved the redundancy problem by retaining only one read pair per mutation/error site (Supplementary Table 4). Different features were extracted from the overlapping read pairs and the non-overlapping read pairs (Supplementary Table 1). All categorical features were expanded using the one-hot encoding method. The hyper-parameters of the random forest model were as follows: (1) the number of decision trees was 100, (2) the maximum tree depth was 50, (3) imbalanced classes were addressed by setting the class weights to “balanced”, and (4) other parameters were left at their default values. Two separate random forest classifiers (one for overlapping read pairs and one for non-overlapping read pairs) were trained on the extracted read pairs.

We validated the performance of the random forest model by cross-validation. For each patient, the labeled read pairs from the 17 other patients were used to train the model, while the patient’s own data were used to test the model (results shown in Supplementary Figure 6). The training data of the random forest model in all the simulation (MRD/recurrence and second primary cancers) also exclude the patient used for generating the simulation data to avoid data leakage. Therefore, the evaluation of *cfTrack* is independent of the training data. As an independent validation set, we used a group of non-small-cell lung cancer patients (8 patients each with 3 samples) with sequential plasma cfDNA samples. The read pairs in these cfDNA samples were labeled in the same manner as described above. Then, these labeled read pairs were used as independent testing data for the random forest model trained by the data generated from the 12 MBC and 6 CRPC patients. On all cross-validation datasets, the random forest model can accurately distinguish sequencing errors from true variants (average AUC = 0.95, 95% confidence interval (CI) = 0.9496-0.9503).

### Simulation of recurrence and MRD detection by tracking clonal somatic mutations in pre-treatment samples

To evaluate the performance of our method, we generated simulation data to mimic patients with MRD/recurrence and patients with complete remission. The patients with MRD/recurrence have tumor content in the post-treatment plasma sample and will show the detection sensitivity; the patients with complete remission have no tumor content in the post-treatment plasma sample and will show the detection specificity. The simulation data were generated from two datasets independently: (1) validation dataset, 27 MBC and 14 CRPC patients and (2) independent dataset, 8 NSCLC patients.

In the validation dataset, only 12 MBC and 6 CRPC patients have two plasma cfDNA samples, so only these patients were used to generate the post-treatment cfDNA samples from the MRD/recurrence patients. Note that these data were also used to generate the training data for the read-level error suppression model. Therefore, to avoid data leakage in the performance evaluation, the MRD/recurrence detection on the validation dataset was performed in a “leave-one-patient-out cross-validation” manner. In other words, for a simulated sample (generated from WES data from a specific patient) in the validation dataset, the random forest models used in the error suppression step were trained on the other 17 patients. In the independent dataset, the 8 NSCLC patients have three plasma cfDNA samples. Only the first two time points of the plasma cfDNA samples were used in the simulation. These data were untouched and independent of the training of the read-level error suppression model, so the error suppression model used on the independent dataset was trained by all training data extracted from the 12 MBC and 6 CRPC patients.

To demonstrate the sensitivity of detection for pre-existing cancer, we generated *in silico* dilution series to simulate patients with MRD/recurrence by mixing the plasma sample collected at the second time point and the matched WBC sample at varying concentrations of cfDNA reads (0.01%, 0.05%, 0.1%, 0.3%, 0.5%, 0.8%, 1%, 3%, 5%, and 8%) using *samtools view* and *samtools merge*. Five independent mixtures were generated at every concentration, at theoretical depths of 200x, 100x or 50x on the WES targeted regions. Since read sampling is random, it is possible that there is no variant supporting read at a given marker, even across all markers. Thus, we removed samples with no variant supporting reads at all personalized markers (checked by *samtools mpileup*). In this simulation, the original matched WBC samples and the original plasma samples at the first time point were used as the WBC samples and the pre-treatment plasma samples, respectively (Figure 2). The *in silico* dilution series represents post-treatment plasma samples from patients with MRD/recurrence. For the validation dataset, we generated the data for each of the 12 MBC and 6 CRPC patients. The theoretical tumor fraction in each sample is calculated as the product of the original tumor fraction in the cfDNA sample and the dilution. The theoretical tumor fraction ranges from 0.001% to 6.114%, with a median of 0.270%. For the independent dataset, we generated the data for each of the 8 NSCLC patients. The theoretical tumor fraction ranges from 0.001% to 1.867%, with a median of 0.103%. The different ranges of the theoretical tumor fractions in the two datasets are caused by differences in the tumor content levels in the original plasma samples. Note that the theoretical tumor fraction usually overestimates the true tumor fraction because of random sampling and the imperfect on-target rate.

**Figure 2.**
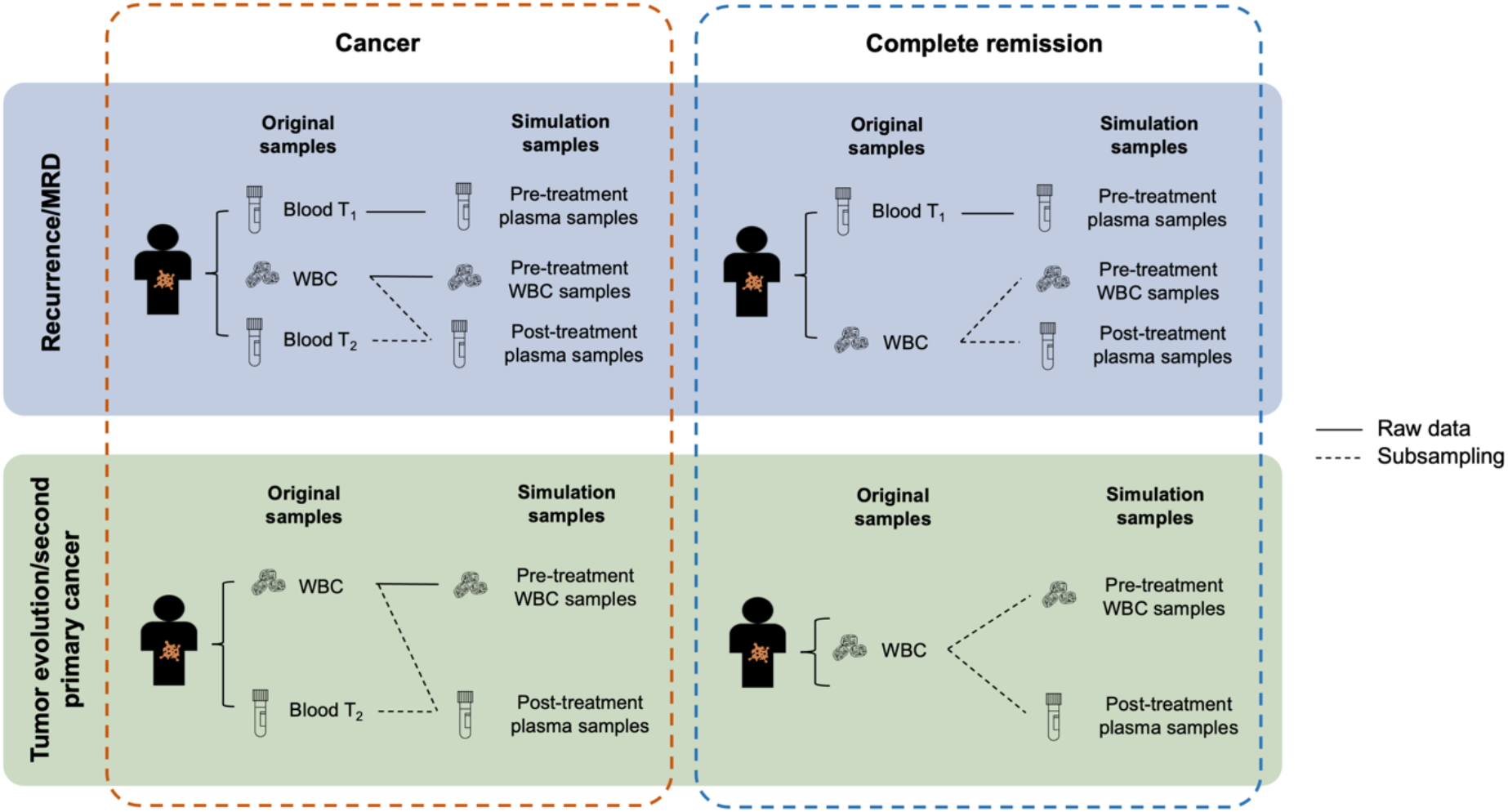
Settings to generate *in silico* spike-in simulation data. The simulation data are generated using WES data taken from (1) 12 MBC and 6 CRPC patients and (2) 8 NSCLC patients. Each patient has an early plasma sample (Blood T_1_), a WBC sample (WBC), and a late plasma sample (Blood T_2_). The three WES datasets from a patient are used directly or mixed to generate the simulation samples. To simulate the scenario of monitoring a patient for MRD or cancer recurrence, each case contains three simulation samples: a pre-treatment plasma sample, a pre-treatment WBC sample, and a post-treatment plasma sample. The raw data from Blood T_1_ are used directly as the pre-treatment plasma sample for all cases. WBC and Blood T_2_ are mixed at specified dilutions to simulate the post-treatment plasma sample. To simulate remission cases, we generate two independent random samplings from the raw WBC data to use as the pre-treatment WBC sample and the post-treatment plasma sample. To simulate the emergence of second primary cancers, each case contains two simulation samples: a pre-treatment WBC sample and a post-treatment plasma sample. The generation of simulation samples for second primary cancer monitoring is the same as for MRD/recurrence monitoring, except that the pre-treatment plasma sample (Blood T_1_) is not used.

To evaluate the specificity of the MRD detection pipeline, we generated the patients with complete remission by subsampling from the original WBC samples. Therefore, these subsamples are expected to have no tumor DNA. For a WBC sample from a cancer patient, five subsamples were generated for each of 200x, 100x, and 50x theoretical depth of the targeted regions. These subsamples represent post-treatment plasma samples from patients without MRD. The original plasma samples at the first time point were used as the pre-treatment plasma samples. Note that only one WBC sample was available for each patient. If the only original WBC sample was directly used as the pre-treatment WBC sample, all data in the post-treatment plasma samples (i.e. subsamples) would have been observed in the pre-treatment WBC sample (i.e. the full original sample), which is impossible in reality. Therefore, we used another subsample of the original WBC samples as the pre-treatment data at a sampling rate of 95% (Figure 2). In this simulation, we preserved some randomness between the WBC samples and the post-treatment plasma samples, which reflects real cases. For the validation dataset, we generated the remission samples for each of the 27 MBC and 14 CRPC patients. For the independent dataset, we generated the remission samples for each of the 8 NSCLC patients.

To avoid potential bias from independently sampling replicates from the same patients, we randomly selected 1 replicate at every dilution (including 0% for remission samples) for every patient to calculate the performance (AUC, sensitivity, and specificity). After the selection, the performance metrics (AUC, sensitivity, and specificity) were evaluated on the MRD/recurrence samples grouped by the tumor fraction with a 0.01% step size and the remission samples (samples with WBC reads only). To provide a robust estimate, we randomly selected samples and calculated the performance 50 times. For the validation dataset, in each random selection, there are 41 simulated remission samples at each depth. At 200x, there are 143 simulated MRD/recurrence samples; at 100x, there are 142 simulated MRD/recurrence samples; at 50x, there are 128 simulated MRD/recurrence samples. For the independent dataset, in each random selection, there are 8 simulated remission samples at each depth. At 200x, there are 68 simulated MRD/recurrence samples; at 100x, there are 65 simulated MRD/recurrence samples; at 50x, there are 56 simulated MRD/recurrence samples.

### Detection of a second primary cancer

A second primary cancer is detected based on a logistic regression model, whose features are the tumor fraction and the number of detected mutations from *cfSNV* in the post-treatment plasma samples. A sample is predicted with second primary cancers if its prediction score is larger than 95% percentile of prediction scores from the remission samples in the training data; otherwise, it is predicted as remission.

### Simulation of second primary cancer detection

Similar to the simulation of recurrence and MRD detection, to evaluate the sensitivity of the method for second primary cancer detection, we generated an *in silico* dilution series by mixing the plasma samples at the second time point and the matched WBC samples from the 12 MBC and 6 CRPC patients at varying concentrations of cfDNA reads (from 1% to 10%: 1%, 3%, 5%, 8%, and 10%) using *samtools view* and *samtools merge*. Since no training and testing of new models is performed in the detection of second primary cancers, this is an independent testing dataset with respect to the detection method. Each spike-in sample contained a total number of randomly sampled reads theoretically equivalent to 200x depth of the targeted regions. Five independent mixtures were generated at every concentration. The tumor fraction in these spike-in samples was quantified by the variant supporting reads at the clonal somatic mutations identified in the original plasma sample. In this simulation, the original matched WBC samples were used as the WBC samples. To demonstrate the specificity of the method, we reused the complete remission samples at 200x generated in the simulation of recurrence and MRD detection. To avoid potential bias from independently sampling replicates from the same patients, we randomly selected 1 replicate at every dilution for every patient to calculate the performance (AUC, sensitivity, and specificity). To provide a robust estimate, we randomly selected samples and calculated the performance 10 times. In each random selection, there were 90 simulated samples from patients with second primary disease and 41 simulated samples from patients with complete remission. To evaluate the performance, after removing the replicates, the simulation data were randomly split into the training set (50%, n = 66) and the testing set (50%, n = 65) ten times. A logistic regression model is trained on the training set and used to predict the presence of a second primary cancer in the testing set. The performance metrics (AUC, sensitivity, and specificity) are evaluated in the testing set on the second primary cancer samples grouped by tumor fraction with a 0.1% step size, but always using the complete set of remission samples.

## Results

### Comprehensive and personalized cancer monitoring using cfDNA

We present a new cancer monitoring method (Figure 1a and Figure 1b), *cfTrack*, that analyzes both pre-existing tumor mutations and newly emerging mutations in post-treatment samples. We developed four major techniques to suppress background noise, generate sample-specific background noise distributions, and achieve comprehensive and sensitive detection of tumor-derived cfDNA. Specifically, we collect a plasma or solid tumor sample and a matched white blood cell (WBC) sample from a patient before the treatment to select markers (i.e., mutations) that are specific to the pre-existing tumor. In the post-treatment plasma samples, *cfTrack* both tracks pre-existing tumor markers and detects new somatic mutations. The techniques in *cfTrack* are summarized below.

#### (1) Integrate all clonal tumor mutations from the pre-treatment samples

Tumor mutations evolve, so any given somatic mutation observed in pre-treatment samples may disappear in post-treatment samples. We perform WES of the pre-treatment samples (solid tumor or plasma samples) and select clonal somatic mutations that appear in all pre-existing cancer cells and have high variant allele frequencies (VAFs) in the cfDNA [4]. Compared to a pre-defined, limited panel of known tumor mutations, the clonal mutations of this specific patient, observed in WES, are more likely to appear in post-treatment samples and are more informative for monitoring the pre-existing tumor [5]. However, when the tumor fraction in cfDNA is very low, WES sequencing at medium depth (100x or 200x) may contain few variant supporting reads at a specific locus. Therefore, to provide a robust mutation-based statistic index in cfDNA, *cfTrack* aggregates variant supporting reads across all clonal somatic mutations (for details, see Methods and Supplementary Figure 1a-b). Specifically, we quantify the tumor fraction using the **i**ntegrated **v**ariant **a**llele **f**requency (**IVAF**), which is the sum of variant supporting reads divided by the sum of all reads at the clonal somatic mutations. Note that a recent publication developed a similar integrative approach using the reads from whole-genome sequencing of cfDNA [11]. Here we show that WES of cfDNA can also be used for ultra-sensitive cancer detection, and given its cost-effectiveness compared to WGS it is more feasible for clinical use.

#### (2) Suppress sequencing errors at the read level with a random forest model

When we integrate tumor reads across a large number of mutation sites to amplify the tumor signal, sequencing errors also accumulate. Therefore, we have developed a method to suppress individual sequencing errors and enhance the signal-to-noise ratio of cancer detection by differentiating the reads containing sequencing errors from those containing true variants. Specifically, this filter is based on a random forest model (for details, see Methods). Previous work has shown that it is possible for machine learning to distinguish true cancer mutations from sequencing artifacts at the read level, and such filters have been used to predict mutations and detect cancer and MRD [11][13]. Unlike these previous works, our method is specifically designed for cfDNA WES data: it incorporates cfDNA fragmentation patterns and read sequence contexts (e.g. nucleotide substitution C>A). Both features are informative to distinguish tumor-derived true mutations and sequencing errors: tumor-derived cfDNA fragments are shorter than non-tumor-derived cfDNA fragments [14][15]; sequencing error rates are associated with nucleotide substitution types [16]. By combining a wide variety of features (Supplementary Table 1), our model automatically discovers feature co-occurrence relationships that are associated with sequencing errors. The random forest model classifies all supporting reads at clonal somatic mutation loci as containing either a true variant or a sequencing error. Only those reads classified as “true variants” are counted as variant supporting reads.

#### (3) Predict recurrence or MRD using sample-specific background noise distribution

To predict whether a patient has recurrence or MRD, we need to compare the estimated tumor fraction with a background noise distribution which represents the error allele fraction in samples from individuals without a tumor. Previous studies usually compared the post-treatment sample of a patient with a cohort of samples from healthy individuals. Because the inter-individual and inter-experimental differences are difficult to model, however, this kind of comparison can introduce prediction bias, and the resulting detection thresholds are difficult to generalize to other experimental protocols. To avoid this limitation, we build the background noise distribution by calculating the IVAF from random genomic positions *in the same sample* (Figure 1b; for details, see Methods). Therefore, this background noise distribution represents the actual error rates observed in this specific sequencing experiment. Recurrence or MRD can then be detected using the empirical *p*-value of the tumor fraction calculated from the pre-existing clonal mutations with respect to the sample-specific background noise distribution (for details, see Methods).

#### (4) Detect tumor evolution by *de novo* identifying newly emerging tumor mutations

Previously described methods for cancer monitoring focus on a predefined mutation panel, which makes it difficult to detect tumor evolution or second primary cancers. Taking advantage of the WES data with broad genome coverage, *cfTrack* performs *de novo* mutation identification to accomplish both. For this we utilize *cfSNV* [12], a method we recently developed for the sensitive and accurate calling of somatic mutations in plasma samples. *cfSNV* specifically accommodates key cfDNA-specific properties, including the low tumor fraction, short and non-randomly fragmented DNA, and heterogeneous tumor content. It addresses the low tumor fraction and tumor heterogeneity in cfDNA by iterative and hierarchical mutation profiling, and ensures a low false-positive rate by multilayer error suppression. Based on the mutation calling results from cfDNA, we can directly detect tumor evolution or the presence of second primary cancers in terms of *de novo* mutations and the corresponding tumor fraction aggregated across mutation sites (for details, see Methods).

### Analytical performance of detecting cancer recurrence and MRD

To evaluate the performance of *cfTrack* on cancer MRD or recurrence, we use the *in silico* method of preparing spike-in simulation data. If a cancer patient has cancer recurrence or MRD, the post-treatment plasma of the patient will contain DNA corresponding to the pre-existing tumor. To simulate the post-treatment plasma samples from the patients with cancer recurrence or MRD, we computationally mix a plasma sample from a cancer patient with a WBC sample from the same patient. The data, with known dilution ratios, can provide a sensitivity/specificity assessment on *cfTrack*.

We generated two sets of *in silico* spike-in simulation data (see Methods): (1) validation dataset, using the WES data from 12 patients with metastatic breast cancer (MBC) and 6 patients with metastatic prostate cancer (castrate-resistant prostate cancer, CRPC) [17] and (2) independent dataset, using the WES data from 8 patients with non-small cell lung cancer (NSCLC). For both datasets, each patient has sequencing data from two plasma samples (collected at two different time points T_1_ and T_2_, with 14∼138 days in between for MBC and CRPC patients, 42 days in between for NSCLC patients), and the matched WBC sample. These patients underwent treatment between T_1_ and T_2_, so we consider the first plasma samples (at T_1_) the “pre-treatment” samples, and the second plasma samples (at T_2_) the “post-treatment” samples. Tens to hundreds of clonal somatic mutations (for MBC and CRPC patients, ranging from 49 to 674 with median 94; for NSCLC patients, ranging from 30 to 1239 with median 63) are found in the pre-treatment samples when compared to their matched WBC samples. We then generate an *in silico* dilution series for each patient by mixing their post-treatment plasma sample with the matched WBC sample at varying fractions (the theoretical tumor fraction ranges from 0.001% to 6.114% with median 0.270% for the validation dataset, from 0.001% to 1.867% with median 0.103% for the independent dataset; for details, see Methods and Figure 2). In addition, we simulate patients who achieved complete remission by subsampling the original WBC samples (the tumor fraction is 0%, for details see Methods and Figure 2). The simulation data are generated at three different depths, 50x, 100x and 200x.

When applying *cfTrack* to the simulated datasets, we observe slightly increased detection performance with increasing sequencing depth (Figure 3a-d and Supplementary Figure 2a-d). This trend is expected because the higher the sequencing depth, the more tumor DNA fragments can be captured. Specifically, on the validation dataset, we achieve an average AUC of 99% (standard deviation (SD) = 1%) when the tumor fraction is ≥ 0.05% at 200x depth (Figure 3a and Supplementary Figure 2a), with 100% average sensitivity (SD = 0%) and 96% average specificity (SD = 1%, Figure 3b and Supplementary Figure 2b). On the independent dataset, we achieve an average AUC of 100% (SD = 0%) when the tumor fraction is ≥ 0.05% at 200x depth (Figure 3c and Supplementary Figure 2c), with 89% average sensitivity (SD = 13%) and 100% average specificity (SD = 0%, Figure 3d and Supplementary Figure 2d). Considering the difference in the sample size and the higher specificity in the independent dataset, the performance on the two simulation datasets is comparable. This indicates that our method can achieve sensitive monitoring using only 200x WES data, offering a cost-effective solution for MRD detection. The detection limit can be further enhanced by increasing the sequencing depth.

**Figure 3.**
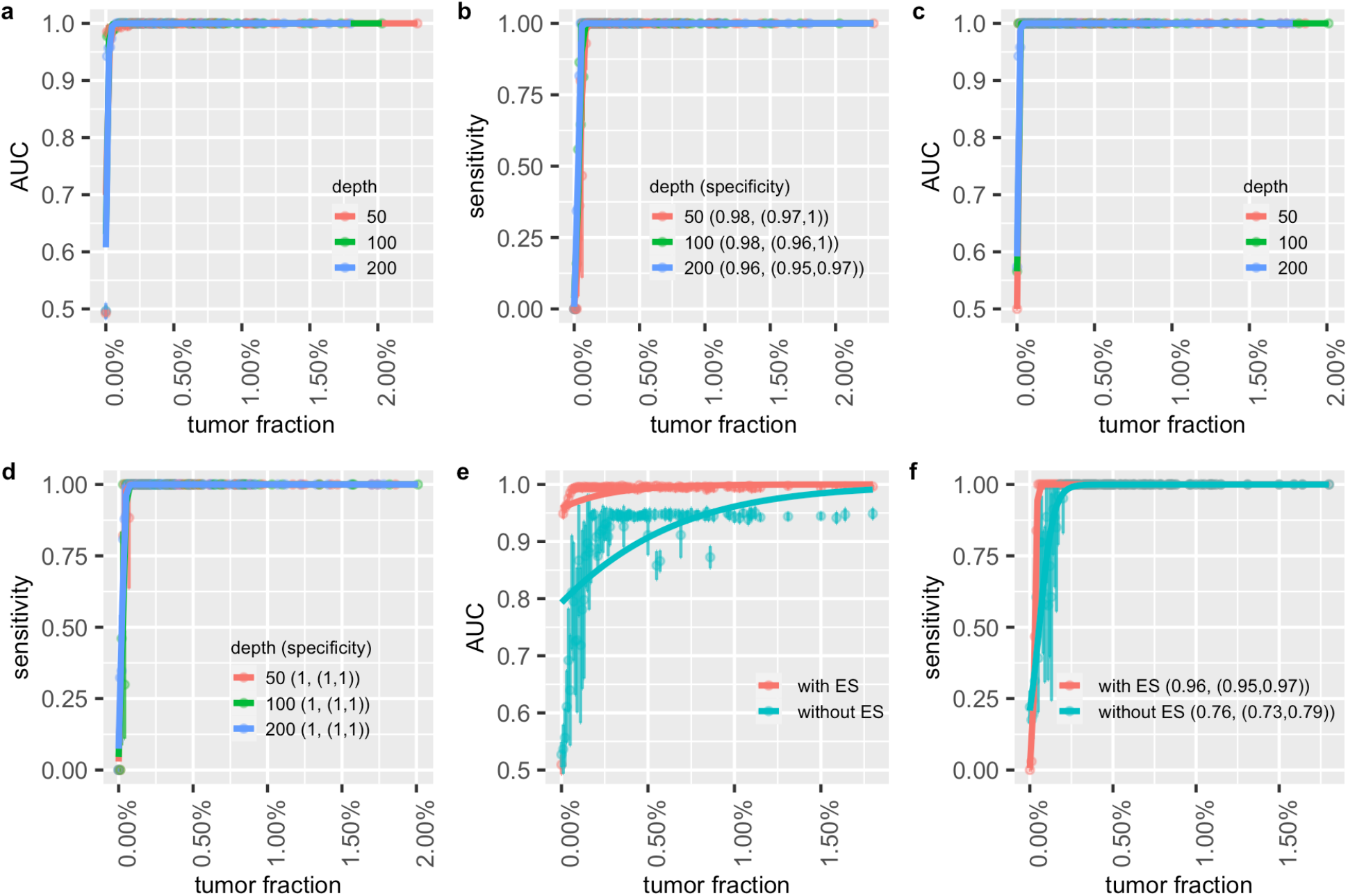
Performance of cancer recurrence and MRD detection using the simulation data. The area under the ROC curve (AUC) of the MRD/recurrence detection on **(a)** the validation dataset and **(c)** the independent dataset with different tumor fractions and sequencing depths. The sensitivity and specificity with different tumor fractions and sequencing depth on **(b)** the validation dataset and **(d)** the independent dataset. Supplementary Figure 2 (a-d) is the zoom-in of (a-d) at low tumor fraction ranging from 0% to 0.2%. **(e)** AUCs of MRD/recurrence detection with and without error suppression (ES) on the validation dataset at 200x depth with different tumor fractions. **(f)** The sensitivity and specificity of MRD/recurrence detection with and without error suppression on the validation dataset at 200x depth with different tumor fractions. In **(a), (c)** and **(e)**, the dots indicate the average AUC, and the vertical bars indicate average *±* SD of the AUC (see Methods). In **(b), (d)** and **(f)**, the dots show the average sensitivity using a cutoff p-value = 0.05 for the background noise distribution; the vertical bars indicate average *±* SD of the sensitivity; the specificity is shown in the legend in the format of (average specificity, (average - SD, average - SD)). The solid lines show the smoothed performance fitted with logit functions.

Our method can achieve the high detection power thanks to three key features: the exome-wide integration of tumor signals, the sample-specific decision threshold, and the read-level error suppression. Read-level error suppression greatly improves the detection power, especially in samples with a low tumor fraction. For example, based on our *in silico* samples with a 0.05% tumor fraction, employing read-level error suppression improved AUC by 35% on the validation dataset (see Figure 3e and 3f) and improved AUC by 40% on the independent dataset (see Supplementary Figure 2e and 2f).

### Analytical performance of detecting second primary cancers

Sensitive monitoring of tumor evolution and newly emerging tumors requires the *de novo* detection of mutations from previously unobserved tumors. Pre-treatment plasma samples and tumor biopsy samples cannot provide sufficient tumor markers for this purpose. In contrast with previous cancer monitoring methods, we can detect *de novo* tumor-derived SNVs in the post-treatment plasma samples, which allows us to identify mutations that come from new tumors. In this section, we specifically evaluate *cfTrack* for the detection of second primary cancers, which depends solely on the detection of emerging tumors.

Detecting a second primary cancer is equivalent to detecting a new tumor without prior knowledge. To simulate this scenario, we generate an *in silico* dilution series from the 12 MBC and 6 CRPC patients by mixing their post-treatment plasma samples with the matched WBC samples [17]. The mixed samples are prepared at varying fractions (the theoretical tumor fraction ranges from 0.111% to 7.680%, with a median of 2.984%; for details, see Methods and Figure 2). For each dilution level, simulation data are generated with a depth of 200x. The samples simulating complete remission are the same as those used for MRD/recurrence detection (in the previous section). Since the detection of a second primary cancer involves no training or testing of new models, this simulation dataset is an independent dataset with respect to the detection method. In this simulation, we do not use the pre-treatment plasma samples, representing the scenario where no pre-existing tumor profile has been observed.

For each pair of simulated plasma and simulated WBC samples, we use *cfSNV* to identify somatic mutations. Then *cfSNV* estimates a tumor fraction from these mutations. We predict a second primary cancer by a logistic regression model using both the tumor fraction and the number of detected mutations as features. We randomly split the samples into a training set (50%) and a testing set (50%). A patient is predicted to have a second primary cancer if they have a large prediction score (≥ 95th percentile of prediction scores from the remission samples in the training set). The AUC is calculated based on the prediction results in the testing sets for all complete remission samples and for the subset of simulation samples with a specific tumor fraction (see Methods). We achieve an average AUC of 88% (SD = 10%) when tumor fraction ≥ 0.2% at 200x depth (Figure 4a), with an average sensitivity of 76% (SD = 23%) and an average specificity of 93% (SD = 5%, Figure 4b). The sensitivity of the methodology is lower for detecting second primary cancers than for detecting recurrence and MRD, because no pre-existing tumor information is available and all novel somatic mutations need to be confirmed. The detection of a novel somatic mutation requires more variant supporting reads than just observing a weak signal at a known locus. Nevertheless, *cfTrack* still achieves high performance in detecting a new tumor. Therefore, *cfTrack* can be used for monitoring tumor evolution and detecting second primary cancers and cancer progression.

**Figure 4.**
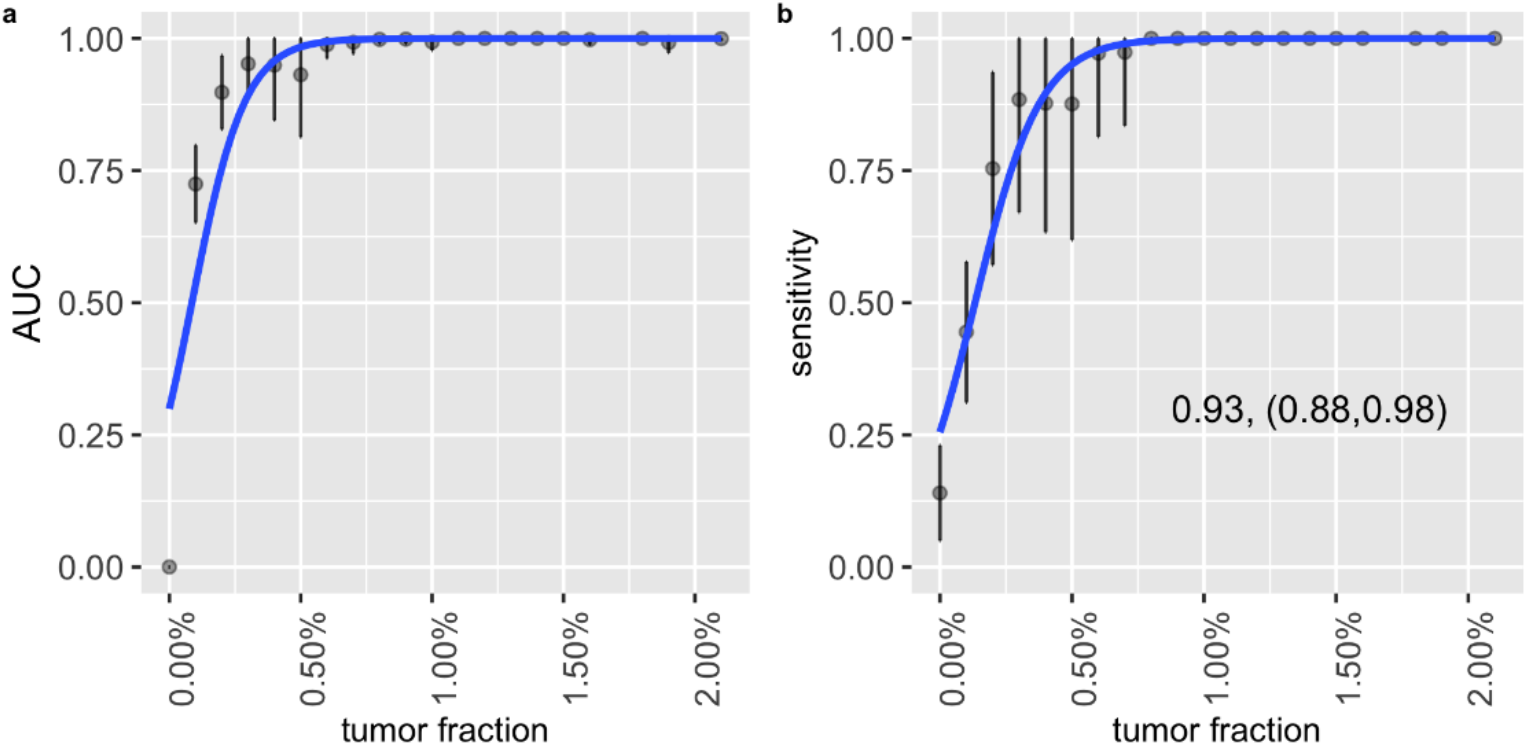
Performance of second primary cancer detection with the simulation data. **(a)** AUC of the *in silico* spike-in samples with different tumor fractions at 200x sequencing depth. The dots indicate the average AUC, and the vertical bars indicate average *±* SD of the AUC (see Methods). **(b)** The sensitivity and specificity in the *in silico* spike-in samples with different tumor fractions at 200x sequencing depth. The dots show the average sensitivity using a cutoff of the 95th percentile of prediction scores from the remission samples in the training data; the vertical bars indicate average *±* SD of the sensitivity; the specificity is shown in the text in the format of (average specificity, (average - SD, average + SD)). The solid lines show the smoothed performance fitted with a logit function.

### Monitoring tumors in cancer patients on treatments through cfDNA

Developments in immunotherapy and targeted therapy have improved the outcomes of cancer patients in recent years [18][19][20]. For example, immunotherapy, which activates a patient’s own immune system to fight cancer, has remarkably improved clinical outcomes in a subset of NSCLC patients [21]. Despite these results, the majority of patients eventually develop resistance and fail to respond to treatment [22][23][24]. Therefore, it is essential to closely monitor the response of patients and quickly recognize when the need for alternative treatment arises. However, since the development of resistance may be associated with tumor evolution [25], this type of monitoring cannot only rely on markers derived from the pre-existing tumor, but requires constant re-evaluation of the tumor profile during treatment. Our WES-based method, which detects mutations from both pre-treatment and treated samples, can comprehensively track a patient’s response.

To test our method in this clinical scenario, we applied our cancer monitoring method to plasma/serum samples (n = 76, 8 serum samples for 4 ovarian cancer patients and 68 plasma samples for other patients) from a cohort of cancer patients (n = 35) who received various treatments. This cohort contains 18 prostate cancer patients [26][27], 8 lung cancer patients, 4 ovarian cancer patients, 3 glioma patients, 1 bladder cancer patient [27], and 1 germ cell cancer patient [27]. All plasma/serum samples were collected when the patients didn’t have complete remission or had recurrence, so tumor content was expected in all samples. After applying our method, tumor-derived DNA was detected in all cfDNA samples except three plasma samples from glioma patients (Supplementary Figure 3). Because the detection of tumor-derived cfDNA is only possible in a very small fraction of glioma patients due to the blood-brain barrier [28], our results were reasonable and consistent with the literature.

Among the 35 patients, 8 NSCLC patients, 4 ovarian cancer patients and 12 prostate cancer patients have at least two plasma/serum samples collected at different time points, between which the patients received treatments. To monitor the tumor changes in these patients, two tumor fractions are calculated separately for the pre-existing tumor mutations (pre-existing tumor fraction) and for the *de novo* tumor mutations (*de novo* tumor fraction) from *cfTrack*. The two tumor fractions allow us to track possible tumor mutations during treatment.

The eight NSCLC patients received anti-PD-1 immunotherapy and their plasma samples were collected from each patient at 0 weeks (baseline), 6 weeks and 12 weeks, measured from the start of treatment. Among these patients, four are “durable responders” whose progression-free survival (PFS) is longer than 18 months; the other four patients are “early progressors” whose PFS is shorter than 6 months (see Supplementary Table 2). In general, we observe a decreasing or low tumor fraction in the durable responders and an elevated tumor fraction in the early progressors (Figure 5a). An unusual example in the sample is early progressor LC-2, whose pre-existing tumor fraction remained at a low level during immunotherapy treatment, while *de novo* tumor fraction increased. This implies a potential clonality change during treatment. In other words, the responding clone might have shrunk while the other clones grew. Existing cancer monitoring methods, which do not consider newly emerging mutations, could not have recognized this tumor growth and would have misled further treatments.

**Figure 5.**
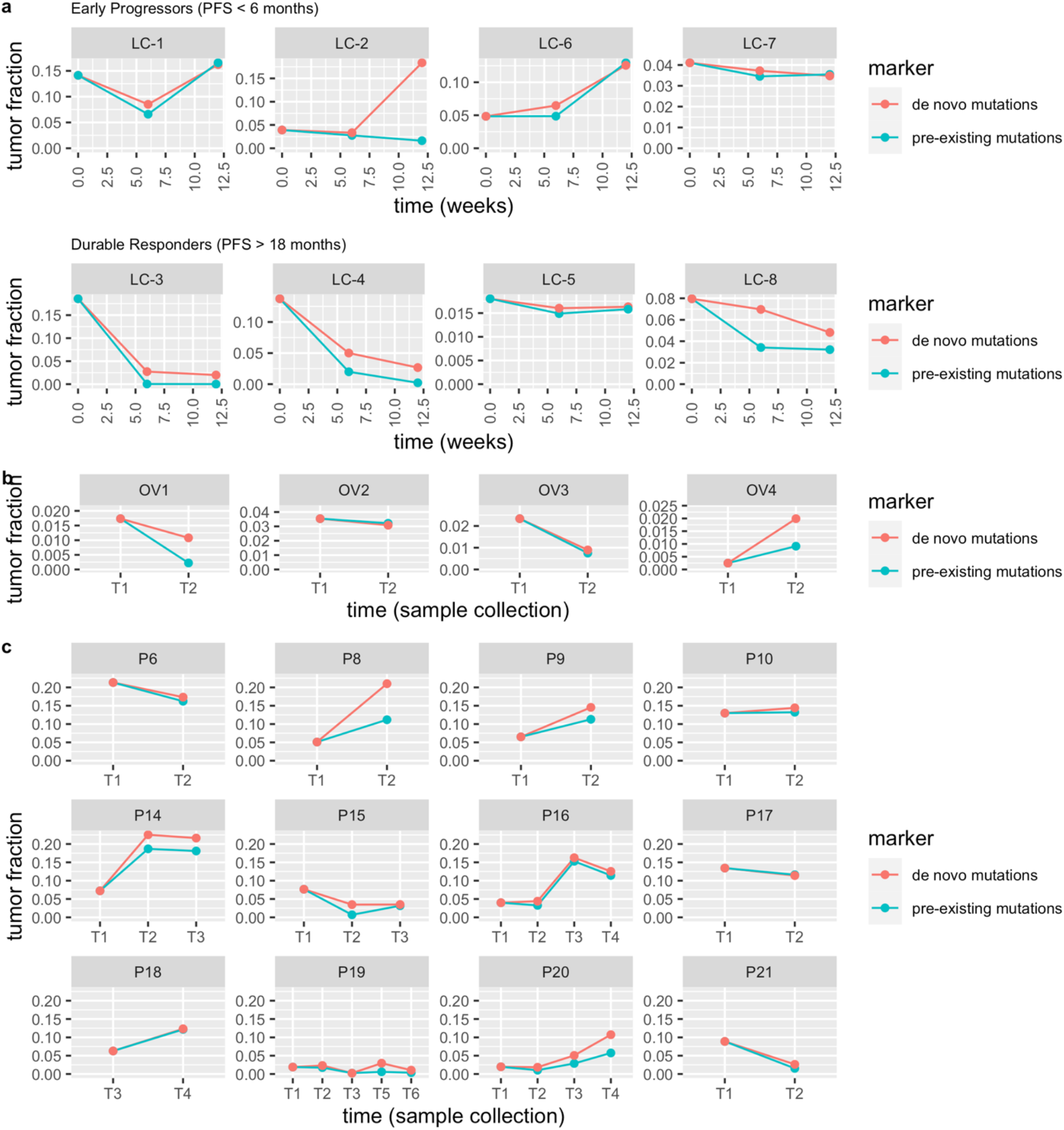
Longitudinal cfDNA monitoring in cancer patients who received treatments. The lines show the tumor fraction in cfDNA during treatment. **(a)** Tumor fraction in plasma samples of 8 NSCLC patients who received anti-PD-1 immunotherapy. **(b)** Tumor fraction in serum samples of 4 ovarian cancer patients. **(c)** Tumor fraction in plasma samples of 12 prostate cancer patients.

The four ovarian cancer patients received chemotherapy (OV1, OV2, and OV3) or chemotherapy and surgery (OV4) between the collection of two serum samples (Supplementary Table 3). At the time of the second collection, patients OV1, OV2, and OV3 underwent surgery. Surgical and pathologic findings demonstrated a moderate treatment effect from chemotherapy. We observed a decrease in both tumor fractions using *cfTrack* (Figure 5b), which indicated a decline in tumor burden. Patient OV4 had a recurrence after chemotherapy and surgery at the time of the second serum collection. Consistently, we observed an increase in both tumor fractions (Figure 5b). Therefore, our results are consistent with the clinical outcomes of these patients.

We also tracked the tumor changes in the 12 prostate cancer patients who received various treatment types during the time between the two plasma collections. During treatment, 9 patients (P8, P9, P10, P14, P15, P16, P18, P19, and P20) had clonal expansion and 3 patients (P6, P17, and P21) had persistent clones [26]. The clonality change can be reflected by the discordance of the two estimated tumor fractions. In general, we observed discordance between the two tumor fractions in the majority of the patients with clonal expansion (Figure 5c). There are no or only minor differences between the two tumor fractions in the patients with relatively stable clones (Figure 5c). These observations are consistent with those from the NSCLC patients.

From the analysis of this heterogeneous cohort of cancer patients with different cancer types and various treatments, we showed that our method can not only closely track the change in tumor fraction, but also detect changes in mutation clonality. The latter is essential for the detection of resistance clones in order to promptly guide subsequent treatments, but it cannot be achieved by existing cancer monitoring methods.

## Discussion

Cancer monitoring is essential to assess the effectiveness of treatment and improve the life quality of cancer patients. Unlike traditional tumor biopsies, cfDNA can provide noninvasive and continuous monitoring of cancer patients, but the very low tumor content of cfDNA remains a major challenge. Most current cfDNA-based methods rely on deeply sequencing a small gene panel to detect the weak tumor signal, but this approach cannot comprehensively cover the patient population or detect evolving tumors. Therefore, we have developed a new cfDNA-based cancer monitoring method that can effectively and sensitively track changes in tumors, detect cancer MRD/recurrence, and identify the presence of a second primary cancer. We present a new computation method for cancer monitoring using cfDNA WES data to overcome the limitations of previous methods. Taking advantage of the wide genome coverage of WES data, *cfTrack* (1) enhances the tumor signal by integrating a large number of clonal tumor mutations identified in pre-treatment samples; (2) suppresses sequencing errors at the read level with an accurate random forest model; (3) builds sample-specific background noise distributions to predict MRD/recurrence, avoiding biases due to inter-individual and inter-experimental variations; and (4) detects tumor evolution and second primary cancers by *de novo* identifying emerging tumor mutations.

Combining these techniques, *cfTrack* achieves sensitive and specific detection of recurrence, MRD and second primary cancers. In detecting recurrence in samples with a 0.05% tumor fraction, *cfTrack* achieved an AUC of 99% (100% sensitivity and 96% specificity) on the validation dataset and an AUC of 100% (89% sensitivity and 100% specificity) on the independent dataset. In detecting second primary cancers in samples with a 0.2% tumor fraction, *cfTrack* yielded an AUC of 88% (76% sensitivity and 93% specificity). Since the performance of the method increases with the sequencing depth, these results can be further improved in practice. As an application, we show that *cfTrack* achieved accurate and comprehensive monitoring of the changes in tumors for patients with different cancer types and undergoing various treatments, which cannot be accomplished by methods focusing only on a small panel of mutations from pre-treatment tumor samples.

This study has its limitations. Firstly, *cfTrack* has only been validated and evaluated using *in silico* spike-in simulation data and on a limited number of cancer patients. To address this limitation, we generated simulation data that mimic real scenarios, including tumor evolution during treatment. For example, simulated plasma samples with tumor content are generated by subsampling the original plasma sample from the second time point, which already contains a different tumor profile compared to the sample at baseline. Nevertheless, we acknowledge that real cases of MRD, recurrence and second primary cancers could be more complicated. Applying *cfTrack* to larger datasets would enable a more comprehensive evaluation and possible optimization of parameters. Secondly, tumor fraction is calculated as an average across all reads for a predefined list of tumor markers. Tumor evolution and tumor heterogeneity could bias the selection of markers, resulting in the absence of important variant supporting reads in the post-treatment cfDNA samples and causing the model to infer a lower tumor fraction. Thirdly, given the medium depth of WES data and the low tumor fraction in the cfDNA samples, *cfTrack* focuses on tracking the overall tumor changes rather than specific clones/subclones. For the same reason, *cfTrack* can detect *de novo* mutations to monitor newly emerging tumors, but it doesn’t guarantee the detection of specific variants directly related to treatment targets.

In this study, for some patients, we use plasma samples to detect the pre-existing tumor mutations, with no need for solid tumor biopsy samples. This is possible as long as the tumor content in plasma samples is sufficient for mutation detection. For patients who receive surgical tumor removal or for patients whose tumor biopsy samples are available, our method can also use a solid tumor sample to identify the pre-existing tumor mutations. However, it is worth noting that a plasma sample may still offer a more comprehensive mutation profile than a biopsy sample [29].

Currently, *cfTrack* utilizes tumor somatic mutations to detect cancer. In a future version, more cancer-specific features in cfDNA can be incorporated. Recent studies have discovered that copy number variations, fragment length, and jagged ends of cfDNA are all associated with tumor-derived cfDNA. In our random forest model, we incorporated the fragment length of the DNA fragments to discriminate true variants from sequencing errors. By integrating other features, we may further empower cancer monitoring to provide actionable information and treatment guidance for patients.

## Data Availability

All data produced in the present study are available upon reasonable request to the authors

## Declaration

### Availability of data and materials

*cfTrack* is implemented in Python and is freely available for academic and research usage through our lab website. The sequence data of the eight NSCLC patients and the four ovarian cancer patients are deposited at the European Genome-phenome Archive (EGA) before publication.

