## Supplementary Materials for "*cfTrack*: Exome-wide mutation analysis of cell-free DNA to simultaneously monitor the full spectrum of cancer treatment outcomes: MRD, recurrence, and evolution"

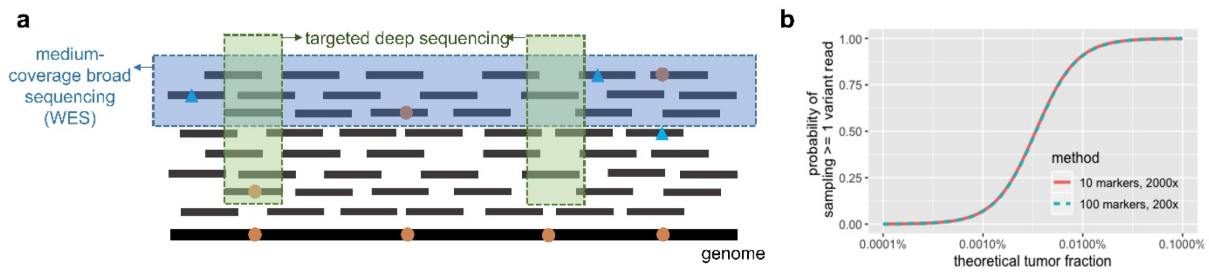


**Supplementary Figure 1. The difference in tumor signal detection between targeted deep sequencing data and medium-coverage broad sequencing data. (a)** Illustration of observed sequencing reads as a sample from the pool of cfDNA. The blue box indicates the observed reads from medium-coverage broad sequencing, while the green box indicates the observed reads from targeted deep sequencing. **(b)** The theoretical detection probability of tracking 10 markers at 2000x and 100 markers at 200x. The probability of sampling >= 1 variant read is determined by a binomial distribution over all markers given a fixed tumor fraction, which is the probability of a read from tumor cells.

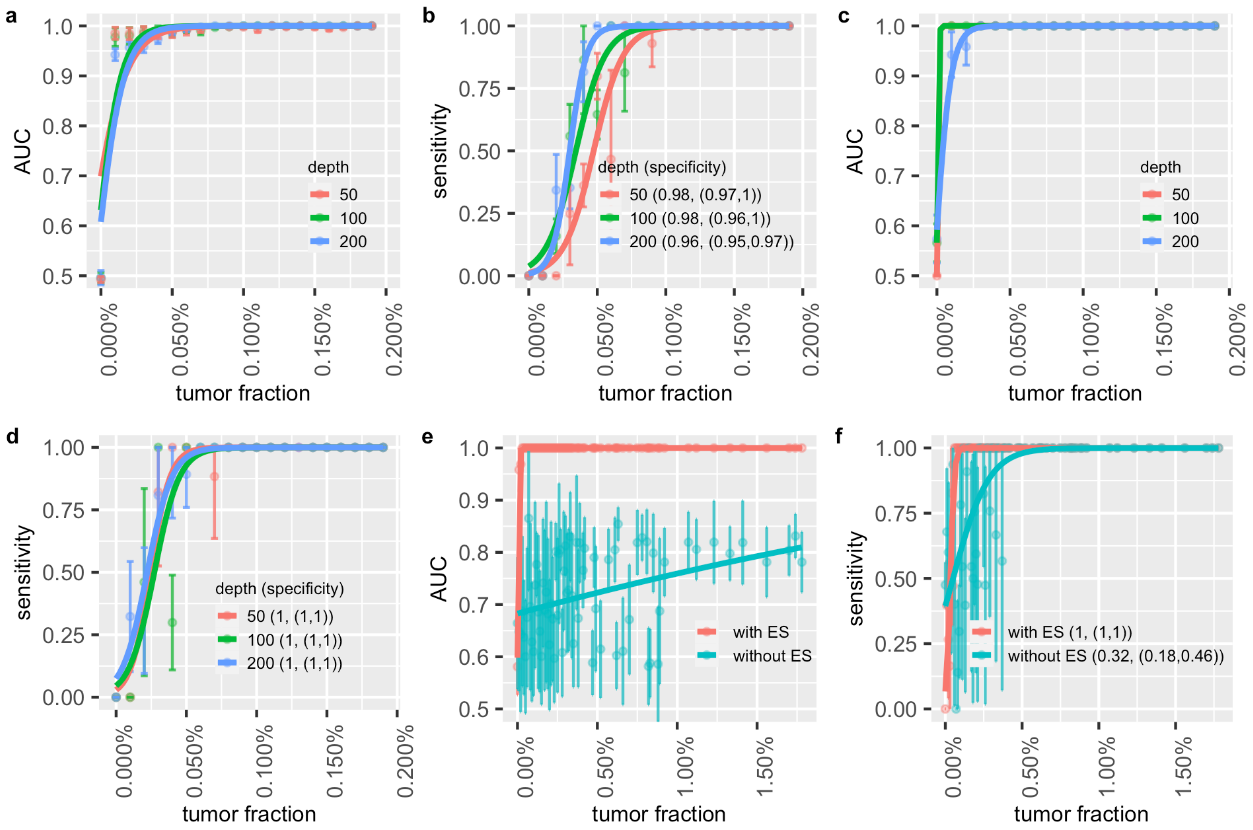


**Supplementary Figure 2. Performance of cancer recurrence and MRD detection using the simulation data. (a-d)** Zoom-in of Figure 3 (a-d). The area under the ROC curve (AUC) of the MRD/recurrence detection on **(a)** the validation dataset and **(c)** the independent dataset with different tumor fractions and sequencing coverage. The sensitivity and specificity with different tumor fractions and sequencing coverage on **(b)** the validation dataset and **(d)** the independent dataset. **(e)** AUCs of MRD/recurrence detection with and without error suppression (ES) on the independent dataset at 200x depth with different tumor fractions. **(f)** The sensitivity and specificity of MRD/recurrence detection with and without error suppression on the independent dataset at 200x depth with different tumor fractions. In **(a)**, **(c)** and **(e)**, the dots indicated the averaged AUC, and the vertical bars indicated average ± SD of the AUC (see Methods). In **(b)**, **(d)** and **(f)**, the dots show the averaged sensitivity using a cutoff p-value = 0.05 of the background noise distribution; the vertical bars indicated average ± SD of the sensitivity; the specificity was shown in the legend in the format of (averaged specificity, (average - SD, average - SD)). The solid lines show the smoothed performance fitted with logit functions.

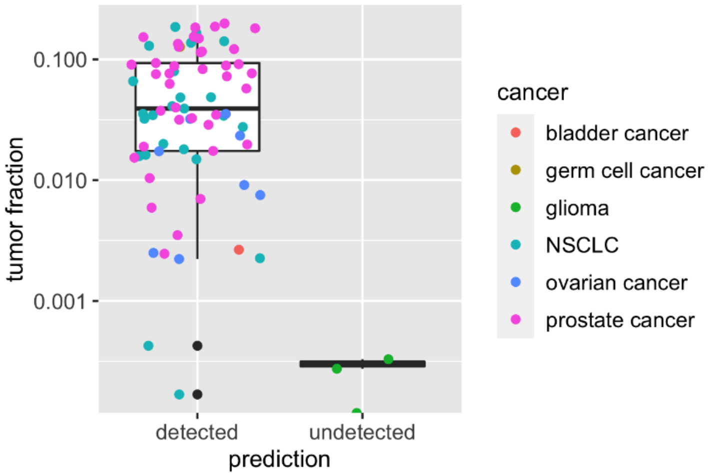


**Supplementary Figure 3. Detection of tumor content in cfDNA from 35 cancer patients.** The dots indicates the cfDNA samples from cancer patients. Each cancer patient contributes 1 - 5 cfDNA samples. The detection of tumor content is based on the cutoff p-value = 0.05 of the background noise distribution.

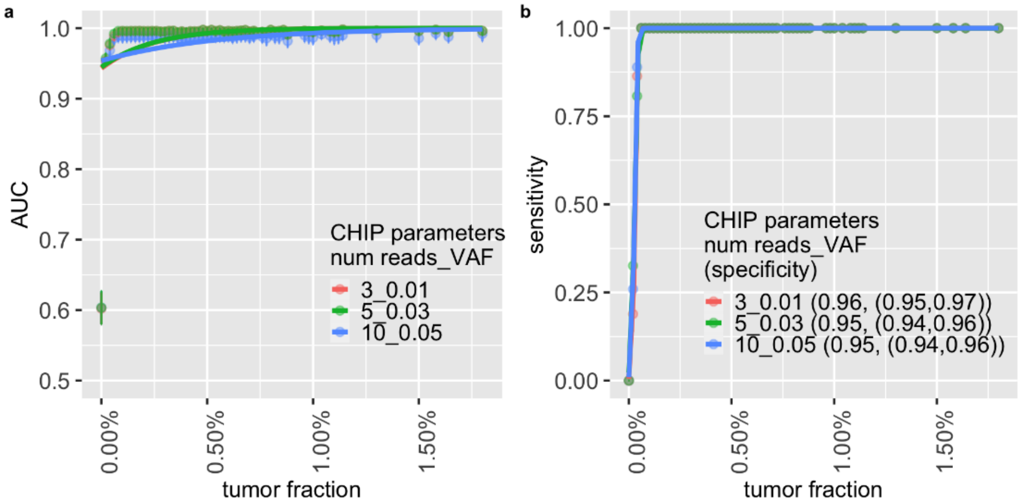


**Supplementary Figure 4. Performance of cancer recurrence and MRD detection using different parameters on the simulation data. (a)** The area under the ROC curve (AUC) of the MRD/recurrence detection on the in silico spike-in samples with different tumor fractions and sequencing coverage. **(b)** The sensitivity and specificity of the in silico spike-in samples with different tumor fractions and sequencing depth. In **(a)** the dots indicate the averaged AUC, and the vertical bars indicate average ± SD of the AUC among the 50 rounds of performance evaluation. In **(b)** the dots show the sensitivity using a cutoff p-value = 0.05 of the background noise distribution; the vertical bars indicate average ± SD of the sensitivity; the specificity was shown in the legend in the format of (averaged specificity, (average - SD, average + SD). The curves in both **(a)** and **(b)** were fitted by logit function. The vertical bars are a little obscure, because the range of the metrics are small and the curves under the three sets of parameters are similar.

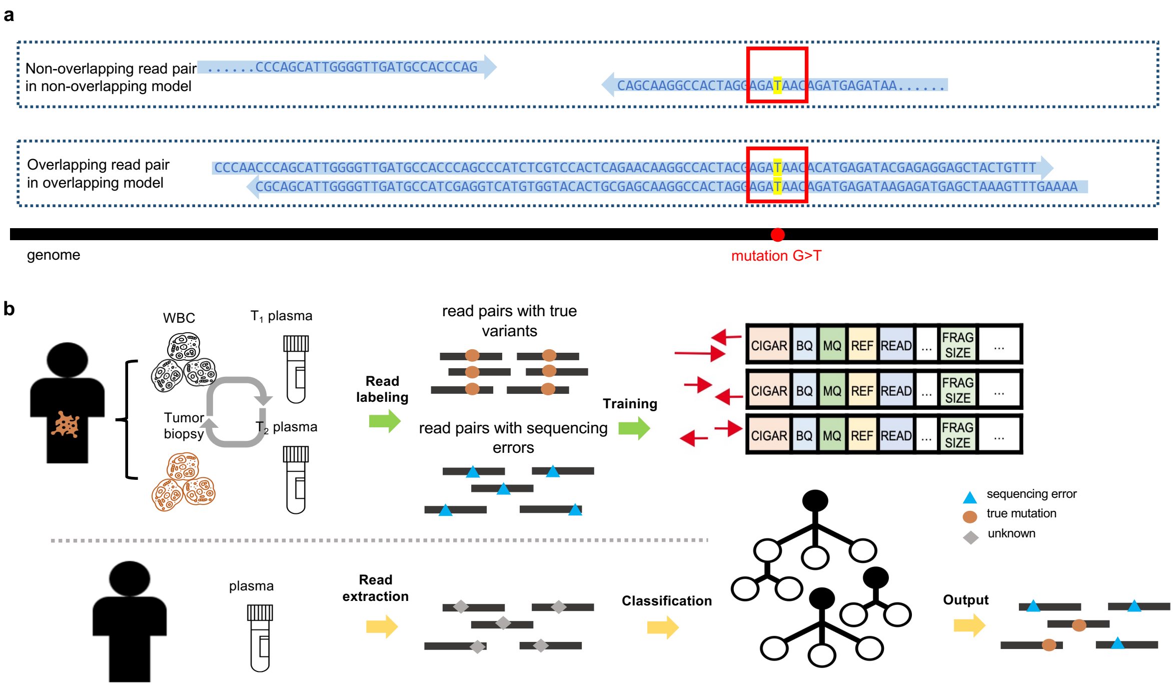


**Supplementary Figure 5. Machine learning model for suppressing sequencing errors. (a)** Read pairs in the overlapping and non-overlapping random forest models. Regarding to a mutation site, there are two types of read pairs: one (non-overlapping read pair) doesn't overlap at the site; the other (overlapping read pair) overlaps at the site. The overlapping read pair naturally contains more information at the mutation site than the non-overlapping read pair. Therefore, two independent random forest model was trained for the overlapping read pair and the non-overlapping read pair. **(b)** Training data extraction and utilization of the random forest model for suppressing sequencing errors at the read level. The upper panel shows the training data extraction workflow (for details, see Methods). True variant positions (somatic and germline mutations) and sequencing error positions are identified by comparing the WBC sample, the tumor biopsy sample and two plasma samples from the same patient. Read pairs with nonreference bases at these identified positions are extracted and labeled "true variants" and "sequencing errors", respectively. Then, various features are extracted from each read pair, and these data are used as training and testing data for the random forest model. The lower panel shows the utilization of the random forest model. Given a post-treatment sample, the features from the read pairs at given loci are extracted from the sequencing data and classified as containing a “sequencing error” or a “true variant”.

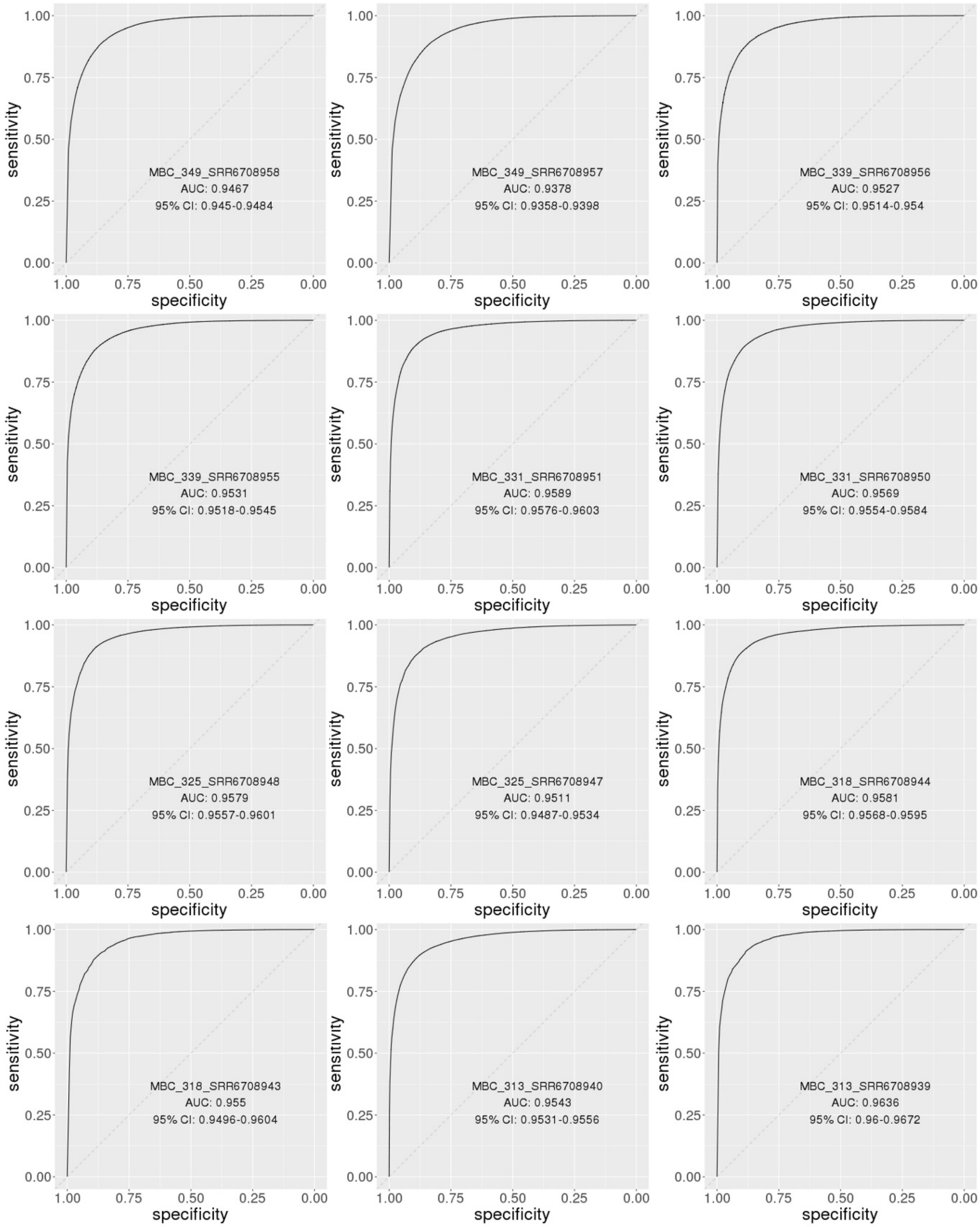

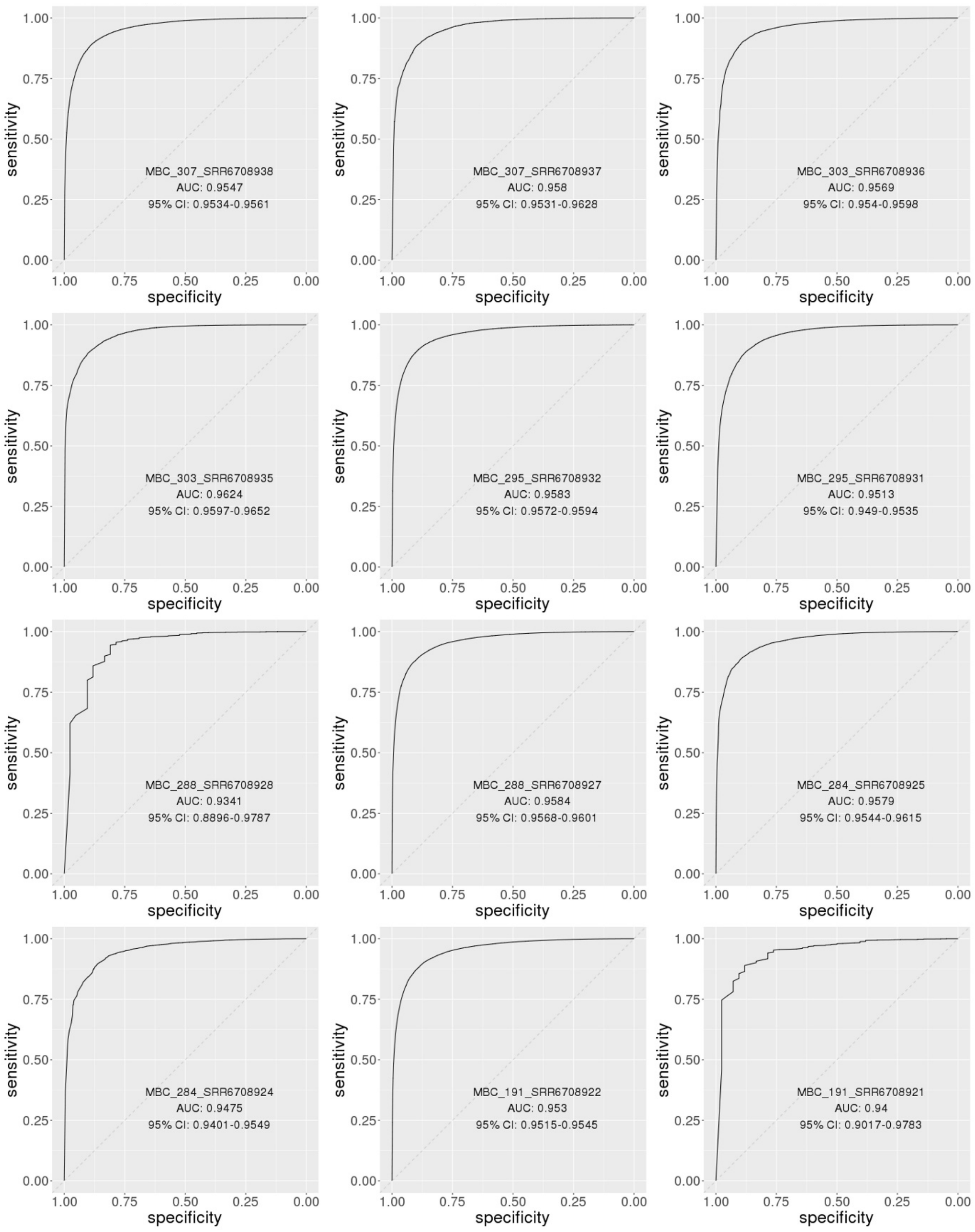
sensitivity
specificity
MBC_307_SRR6708938
AUC: 0.9547
95% CI: 0.9534-0.9561
MBC_307_SRR6708937
AUC: 0.958
95% CI: 0.9531-0.9628
MBC_303_SRR6708936
AUC: 0.9569
95% CI: 0.954-0.9598
MBC_303_SRR6708935
AUC: 0.9624
95% CI: 0.9597-0.9652
MBC_295_SRR6708932
AUC: 0.9583
95% CI: 0.9572-0.9594
MBC_295_SRR6708931
AUC: 0.9513
95% CI: 0.949-0.9535
MBC_288_SRR6708928
AUC: 0.9341
95% CI: 0.8896-0.9787
MBC_288_SRR6708927
AUC: 0.9584
95% CI: 0.9568-0.9601
MBC_284_SRR6708925
AUC: 0.9579
95% CI: 0.9544-0.9615
MBC_284_SRR6708924
AUC: 0.9475
95% CI: 0.9401-0.9549
MBC_191_SRR6708922
AUC: 0.953
95% CI: 0.9515-0.9545
MBC_191_SRR6708921
AUC: 0.94
95% CI: 0.9017-0.9783

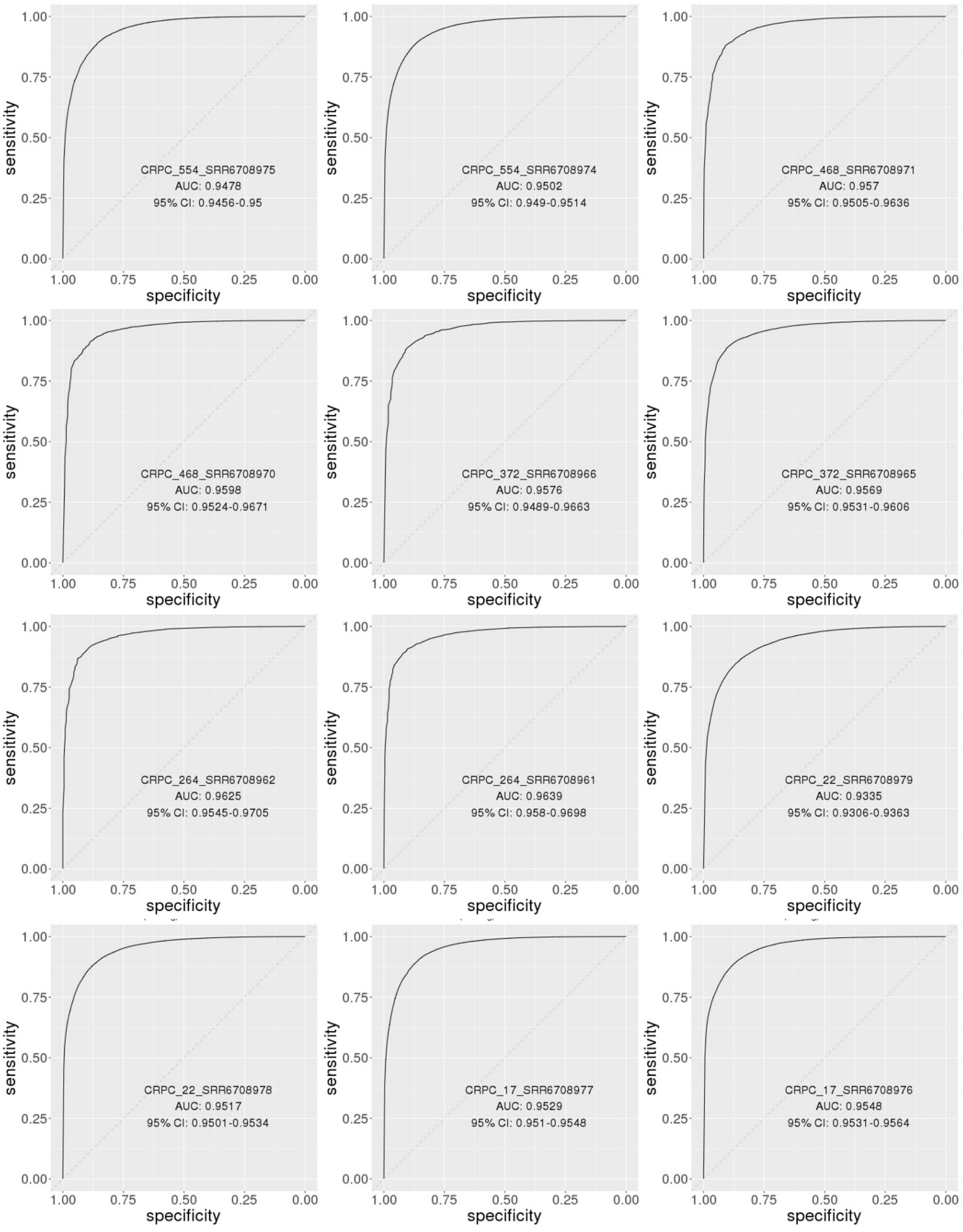


**Supplementary Figure 6. Performance of the random forest model in the 36 cross-validation sets.** The receiver operating characteristic (ROC) curve of the random forest model in the 36 cross-validation sets.

| Feature description | Feature type | Nonoverlapping model | Overlapping model |
|---|---|---|---|
| base call in a 7-bp window centered on the query site from the read | categorical | Yes | Yes |
| base quality in a 7-bp window centered on the query site from the read | numerical | Yes | Yes |
| CIGAR information in a 7-bp window centered on the query site from the read | categorical | Yes | Yes |
| occurrence CIGAR operators from the read | Boolean | Yes | Yes |
| mapping quality of the read pair | numerical | Yes | Yes |
| distance to the nearest indel on the read pair | numerical | Yes | Yes |
| whether the query site was contained in a homopolymer with a size >= 5 | Boolean | Yes | Yes |
| insertion sizes of the read pair | numerical | Yes | Yes |
| genome sequence in a 7-bp window centered on the query site | categorical | Yes | Yes |
| mapping flags of the read and the mate | categorical | Yes | Yes |
| base call in a 7-bp window centered on the query site from the mate | categorical | No | Yes |
| base quality in a 7-bp window centered on the query site from the mate | numerical | No | Yes |
| CIGAR information in a 7-bp window centered on the query site from the mate | categorical | No | Yes |

**Supplementary Table 1. Extracted features from read pairs for the random forest models.** The column “nonoverlapping model” indicates which features are used in the random forest model to filter nonoverlapping read pairs. The column “overlapping model” indicates which features are used in the model for overlapping read pairs.

| Patient ID | PFS (days) | Group |
|---|---|---|
| LC-1 | 133 | Early Progressor |
| LC-2 | 131 | Early Progressor |
| LC-3 | 812 | Durable Responder |
| LC-4 | 951 | Durable Responder |
| LC-5 | 917 | Durable Responder |
| LC-6 | 105 | Early Progressor |
| LC-7 | 84 | Early Progressor |
| LC-8 | > 1825 | Durable Responder |

**Supplementary Table 2. Progression-free survival (PFS) of the 8 non-small-cell lung cancer patients.**

| patient | 1st serum | 2nd serum | Time in between (months) | Notes for chemotherapy |
|---|---|---|---|---|
| OV1 | pre-chemotherapy | post-chemotherapy and pre-surgery | 3.37 | Moderate treatment effect |
| OV2 | pre-chemotherapy | post-chemotherapy and pre-surgery | 2.60 | Moderate treatment effect |
| OV3 | pre-chemotherapy | post-chemotherapy and pre-surgery | 2.60 | Moderate treatment effect |
| OV4 | post-chemotherapy and pre-surgery | post-surgery and chemotherapy recurrence | 9.57 | |

**Supplementary Table 3. Clinical information of the 8 serum samples from the 4 ovarian cancer patients at UCLA.**

| ID | | Error | | Variant | |
|---|---|---|---|---|---|
| Patient ID | Sample ID | No Overlap | Overlap | No Overlap | Overlap |
| CRPC_17 | SRR6708976 | 15680 | 151 | 28570 | 24730 |
| CRPC_17 | SRR6708977 | 10789 | 1358 | 28185 | 25619 |
| CRPC_22 | SRR6708978 | 16282 | 144 | 28622 | 24474 |
| CRPC_22 | SRR6708979 | 6927 | 97 | 27933 | 23840 |
| CRPC_264 | SRR6708961 | 509 | 225 | 27052 | 24457 |
| CRPC_264 | SRR6708962 | 319 | 99 | 27122 | 23835 |
| CRPC_372 | SRR6708965 | 1646 | 584 | 26069 | 23315 |
| CRPC_372 | SRR6708966 | 396 | 92 | 27065 | 24682 |
| CRPC_468 | SRR6708970 | 504 | 198 | 32441 | 29473 |
| CRPC_468 | SRR6708971 | 510 | 239 | 31484 | 27138 |
| CRPC_554 | SRR6708974 | 45617 | 845 | 33894 | 30638 |
| CRPC_554 | SRR6708975 | 9434 | 435 | 33687 | 30218 |
| MBC_191 | SRR6708921 | 39 | 3 | 33692 | 30424 |
| MBC_191 | SRR6708922 | 12953 | 5129 | 32565 | 28987 |
| MBC_284 | SRR6708924 | 594 | 124 | 32020 | 26885 |
| MBC_284 | SRR6708925 | 1726 | 612 | 32400 | 29515 |
| MBC_288 | SRR6708927 | 9658 | 3366 | 32654 | 29815 |
| MBC_288 | SRR6708928 | 38 | 4 | 32852 | 28932 |
| MBC_295 | SRR6708931 | 8895 | 370 | 33720 | 28268 |
| MBC_295 | SRR6708932 | 34007 | 10810 | 32476 | 27214 |
| MBC_303 | SRR6708935 | 3353 | 117 | 34204 | 31599 |
| MBC_303 | SRR6708936 | 3144 | 1126 | 31067 | 26469 |
| MBC_307 | SRR6708937 | 1383 | 40 | 33354 | 29336 |
| MBC_307 | SRR6708938 | 21055 | 5974 | 32663 | 29378 |
| MBC_313 | SRR6708939 | 2292 | 70 | 33820 | 30856 |
| MBC_313 | SRR6708940 | 21503 | 8790 | 31725 | 28822 |
| MBC_318 | SRR6708943 | 1330 | 51 | 33391 | 30548 |
| MBC_318 | SRR6708944 | 16734 | 5850 | 32684 | 29834 |
| MBC_325 | SRR6708947 | 5099 | 2431 | 29623 | 27158 |

| | | | | | |
|---|---|---|---|---|---|
| MBC_325 | SRR6708948 | 6120 | 1521 | 32499 | 29882 |
| MBC_331 | SRR6708950 | 14813 | 4030 | 32628 | 29610 |
| MBC_331 | SRR6708951 | 17448 | 5969 | 32055 | 28088 |
| MBC_339 | SRR6708955 | 28526 | 1651 | 34122 | 30393 |
| MBC_339 | SRR6708956 | 28212 | 1446 | 34088 | 29869 |
| MBC_349 | SRR6708957 | 17462 | 532 | 33951 | 31605 |
| MBC_349 | SRR6708958 | 19876 | 436 | 34059 | 31567 |
| LC-1 | LC-1_12-week | 1 | 0 | 34188 | 34003 |
| LC-1 | LC-1_00-week | 0 | 1 | 32863 | 33395 |
| LC-1 | LC-1_06-week | 4 | 2 | 34186 | 34068 |
| LC-2 | LC-2_00-week | 48627 | 19237 | 34227 | 34139 |
| LC-2 | LC-2_06-week | 38567 | 12011 | 33316 | 33900 |
| LC-2 | LC-2_12-week | 54542 | 7655 | 34522 | 33087 |
| LC-3 | LC-3_00-week | 24672 | 9970 | 32821 | 33588 |
| LC-3 | LC-3_06-week | 19354 | 6022 | 33109 | 33642 |
| LC-3 | LC-3_12-week | 24949 | 5439 | 34582 | 34155 |
| LC-4 | LC-4_00-week | 34884 | 12337 | 32299 | 33056 |
| LC-4 | LC-4_06-week | 38017 | 12905 | 33568 | 33659 |
| LC-4 | LC-4_12-week | 40591 | 12438 | 33622 | 33614 |
| LC-5 | LC-5_00-week | 24 | 11 | 33873 | 33887 |
| LC-5 | LC-5_06-week | 46 | 9 | 33248 | 33635 |
| LC-5 | LC-5_12-week | 22 | 9 | 33244 | 33738 |
| LC-6 | LC-6_00-week | 1071 | 422 | 31756 | 33222 |
| LC-6 | LC-6_06-week | 7433 | 2075 | 34431 | 34373 |
| LC-6 | LC-6_12-week | 13020 | 2695 | 34688 | 34389 |
| LC-8 | LC-8_00-week | 36161 | 12359 | 30874 | 30758 |
| LC-8 | LC-8_06-week | 22368 | 9583 | 30620 | 30743 |
| LC-8 | LC-8_12-week | 23331 | 10328 | 30772 | 30731 |
| LC-7 | LC-7_00-week | 9345 | 4080 | 34467 | 34522 |
| LC-7 | LC-7_06-week | 28379 | 19286 | 34878 | 34570 |
| LC-7 | LC-7_12-week | 17077 | 14692 | 33156 | 33922 |

**Supplementary Table 4. Sample IDs and the number of labeled read pairs for training and testing the random forest model.** For the MBC patients and CRPC patients, the patient IDs follow the naming convention in **Error! Reference source not found.**, while the sample IDs are the SRA accession IDs of the sample.
